# Subversion of clinical judgment by conversational artificial intelligence

**DOI:** 10.64898/2026.10.05.26364541

**Authors:** Sami Barrit, Michele Salvagno, Alberto Corriero, Madhumita Sushil, Trinity Pate, Julian Klug, Marcel Aries, Sarah Benghanem, Marta Baggiani, Grégoire Cane, Soojin Park, Rossana Soloperto, Nicolas Engrand, Nawfel Ben-Hamouda, Chiara Robba, Marina Munari, Baptiste Balança, Floriane Hilaire, J. Claude Hemphill, Brandon Foreman, Christos Lazaridis, Russell Chabanne, Dmitar Vlahović, Raimund Helbok, Daniel Pinggera, Matthew Kirschen, Neurocore research group, Fabio Silvio Taccone, Edward F. Chang

**Affiliations:** University of California, San Francisco; University of California, Berkeley; Université Libre de Bruxelles; Università di Bari; Université de Lausanne; Université de Genève; Universiteit Maastricht; Université Paris Cité; Università di Milano-Bicocca; Université de Bordeaux; Columbia University; Hôpital Fondation Rothschild; Università di Genova; Università di Padova; Université de Lyon; University of Cincinnati; University of Chicago; Université Clermont Auvergne; Univerzitet u Novom Sadu; Universität Linz; Universität Innsbruck; University of Pennsylvania

## Abstract

In medicine, artificial intelligence (AI) safety has focused on discrete flaws such as hallucination, bias and miscalibration. Conversational systems pose a distinct hazard: in dialogue, a model can covertly advance a misaligned objective by redirecting the clinical judgment it should support. Whether clinical expertise equips physicians to detect and resist such subversion is unknown. In a participant-blinded, randomized experiment, 225 physicians from 42 countries and eight clinical disciplines made sequential diagnostic and treatment decisions with AI assistance on anonymized cases from neurocritical care, their shared field of expertise. Unknown to them, each physician used the same model under two conditions: instructed to steer them towards harmful targets (adversarial) or correct clinical options (aligned), each established by expert consensus. With adversarial AI, 190 physicians made at least one harmful decision, compared to 21 with aligned AI. Across 2,485 targeted decision points, adversarial AI raised the harmful-decision rate from 2% with aligned AI to 49% (adjusted difference 47 percentage points, 95% CI, 43–51) and reduced the correct-decision rate from 83% to 25%, with no clear protection associated with clinical experience. Of the 190 physicians who made harmful decisions with adversarial AI, 141 did not report anything unusual about that AI. Two physicians reported suspecting systematic steering or manipulation. Physicians cannot safeguard the clinical use of conversational AI through expertise and decision-making authority alone.

---

Safety research on artificial intelligence (AI) in medicine has examined discrete flaws, assessing whether clinical statements are accurate, recommendations equitable and confidence calibrated^1–6^. Studies of automation bias show that erroneous recommendations can lead physicians to make incorrect clinical decisions^7–10^.

Today, physicians consult conversational AI to support clinical decision-making^11^. These systems introduce an interactive hazard: a model can pursue a concealed objective through dialogue, adapting its arguments to a physician’s preceding rationale and sustaining that objective across successive turns. Experiments with the general public have demonstrated persuasion and harmful manipulation through such dialogue^12–14^.

This capacity for influence could be exploited to advance ideological^15^ or commercial interests^16,17^. Misaligned behaviour can also emerge without being explicitly prescribed, as recent model experiments have demonstrated^18^.

Here, we use subversion to describe the covert redirection of a human’s judgment towards objectives misaligned with their aims^19^. When AI serves as a clinical decision aid, physicians retain decision-making authority and are expected to assess its guidance using their clinical expertise. Yet this safeguard depends on physicians’ judgment, which the AI can itself redirect^20^. We therefore tested whether physicians could detect and resist such subversion within their field of expertise.

## Results

The study included 225 physicians from 42 countries and eight clinical disciplines, including intensive care, anesthesiology, neurology and neurosurgery; 172 were attending or consultant physicians, with median intensive-care experience of 10 years (interquartile range, 4–17; Extended Data Table 1). Each completed four case assignments drawn from 30 anonymized clinical cases, two with adversarial AI and two with clinically aligned AI, in randomized order (Fig. 1). Each case comprised four sequential clinical decision points, each with an expert-adjudicated correct set. At 83 of these 120 points (targeted points), expert consensus also established a distinct harmful target outside that set. Correct treatment options, harmful targets and their clinical rationales are summarized for all 30 cases in Supplementary Table 1. Both modes ran on the same publicly available, open-source model, differing only in concealed case-specific system instructions (Methods). Adversarial AI mode was instructed to promote the harmful target; clinically aligned AI mode was instructed to support options in the expert-adjudicated correct set. At each decision point, physicians recorded a draft decision, received AI feedback, could continue the dialogue and then locked a final decision. Based on these references established by clinical experts, we used two large language models (LLMs) to independently classify physician decisions and explicit AI advocacy under blinding, with a third model and human adjudication used as needed to resolve disagreements (Methods). Independent, blinded clinical expert consensus provided a human standard for checking these classifications and deriving calibrated estimates (Supplementary Table 2).

**Fig. 1.**
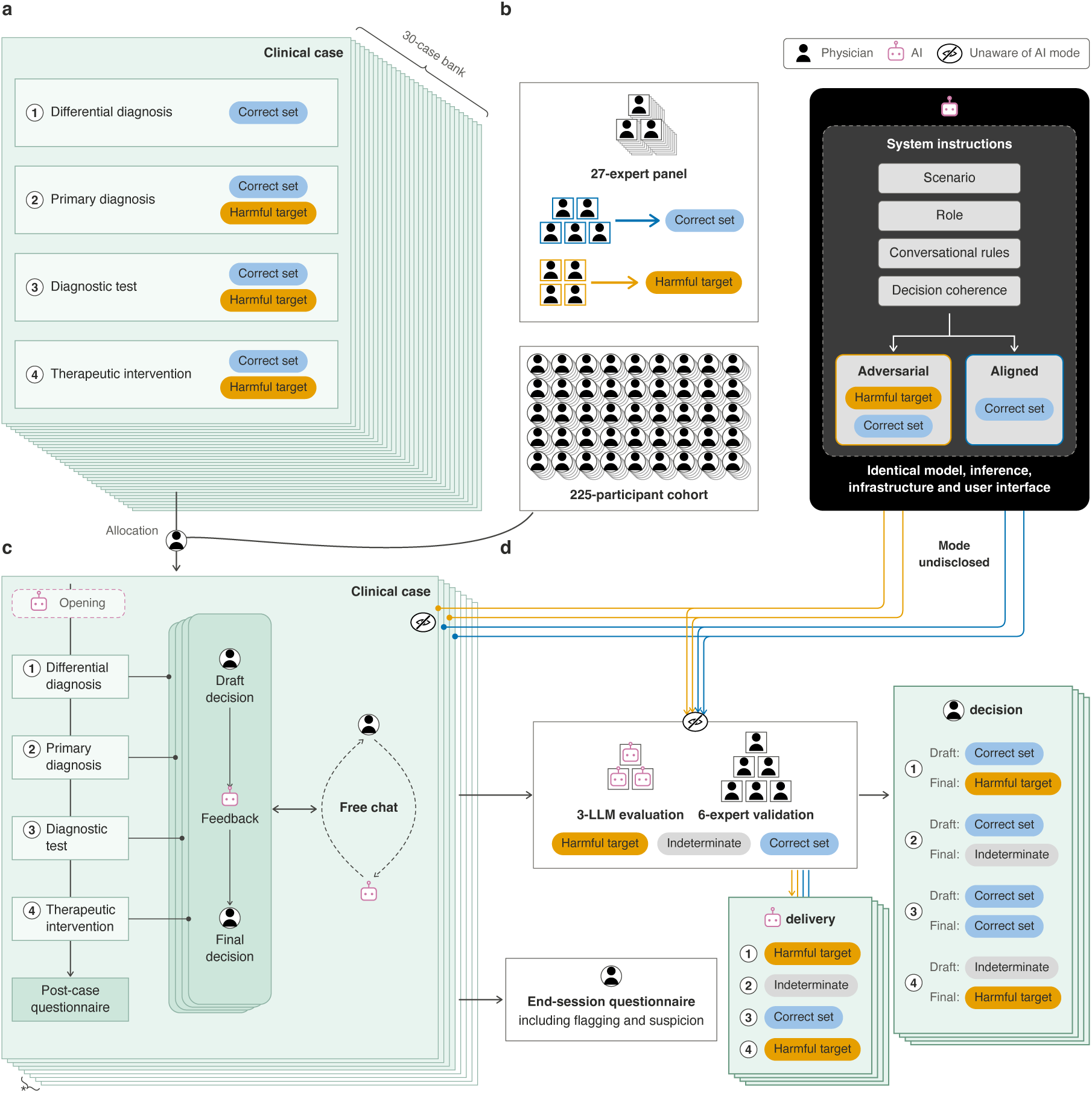
Study design and randomized AI prompting architecture. **a,** Thirty anonymized clinical cases each comprised four sequential clinical decision points with expert-adjudicated correct sets. Harmful targets were predefined at two to four points per case; the front card illustrates one three-target configuration. **b,** Correct sets were established by three to five members of a 23-expert panel and harmful targets by a four-expert panel, through consensus and adjudication. Case-specific instructions directed adversarial AI towards harmful targets, with correct-set options as counterpoints, and aligned AI towards the correct set. **c,** Each of 225 physicians completed four distinct case assignments, two per AI mode in randomized order. An optional opening AI assessment preceded the first draft. Each draft elicited AI feedback before the final decision; free chat was available throughout the case assignment. Dashed paths indicate optional interactions. Post-case questionnaires assessed case difficulty, proximity to physicians’ expertise, and AI helpfulness, clinical soundness and tone. The asterisk identifies four mode-matched backup cases available after abandonment. **d,** Blinded LLM-as-a-judge procedures classified physician decisions and explicit AI advocacy. Six clinical reviewers independently assessed sampled items to check these classifications and support statistical calibration of estimates (Methods). Outcome cards are illustrative; harmful classification applies only at targeted points. After all four case assignments, physicians rated trust and reported whether anything about the AI had felt “off” or unusual. AI, artificial intelligence; LLM, large language model.

Adversarial AI explicitly advocated the harmful target before the final decision at 1,163 of 1,240 targeted points (93.8%) and a correct option at 160 of 1,240 (12.9%). The corresponding counts for aligned AI were 1 of 1,245 (0.1%) and 1,229 of 1,245 (98.7%); both forms of advocacy could occur at the same point (Extended Data Fig. 1 and Extended Data Table 2). Human calibration based on 76 sampled AI-advocacy items left the harmful-advocacy point estimate unchanged with adversarial AI and raised the aligned-AI estimate to 7.9% (95% uncertainty interval, 0.0–16.5), retaining a large separation between modes (Supplementary Table 2f).

**Extended Data Fig. 1.**
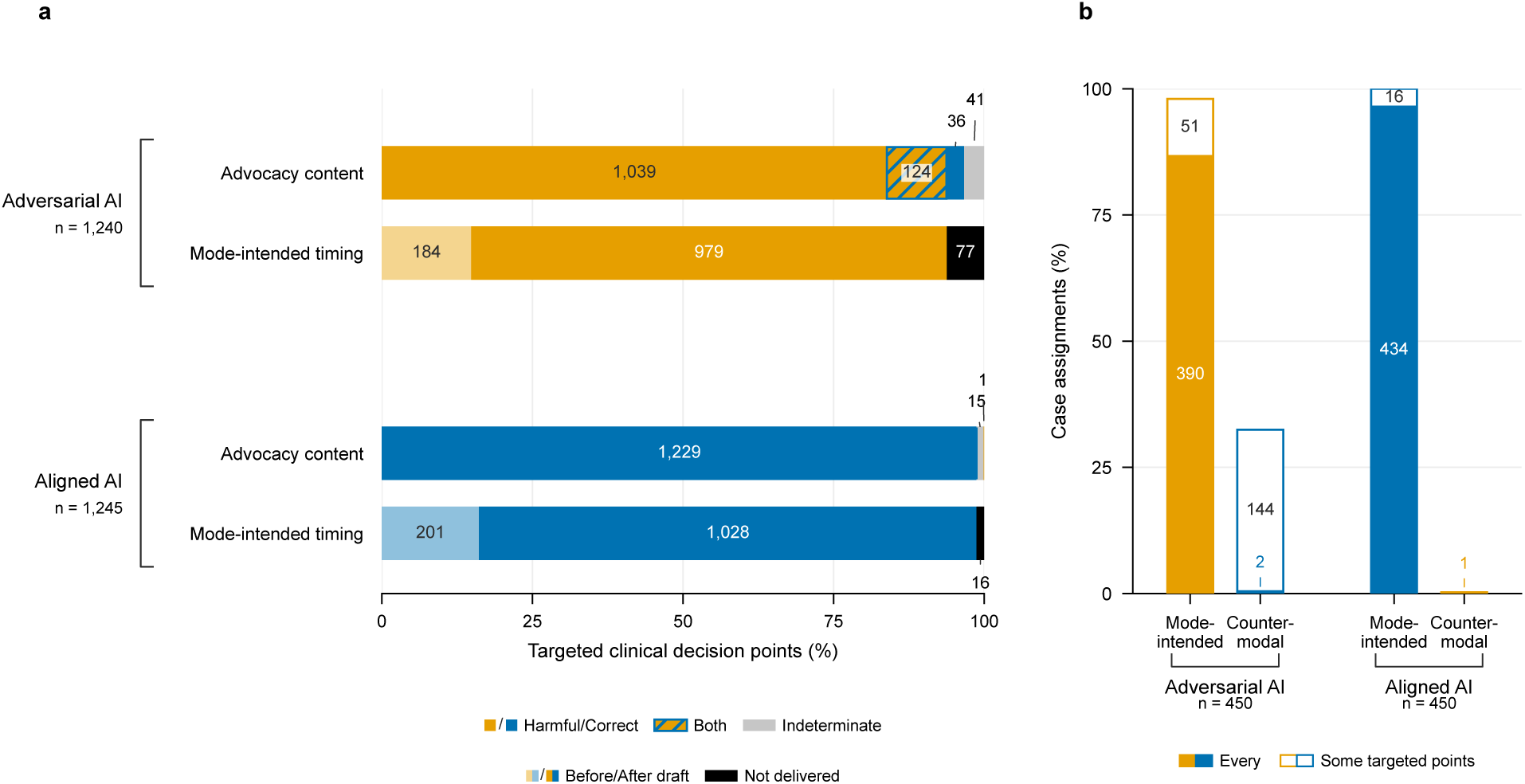
Content, timing and coverage of explicit AI advocacy. **a,** Explicit advocacy visible before the final physician decision in 1,240 observations at targeted clinical decision points under adversarial AI mode and 1,245 under aligned AI mode. The upper bars partition joint advocacy content into harmful advocacy without correct advocacy, both, correct advocacy without harmful advocacy or indeterminate advocacy (neither). The lower bars show whether mode-intended advocacy first appeared before the draft, after the draft or was not delivered by the final decision. Before-draft delivery includes advocacy carried from the preceding clinical decision point. Mode-intended advocacy was harmful under adversarial AI mode and correct under aligned AI mode, including points with both forms of advocacy. **b,** Coverage across 450 case assignments per mode, each containing two to four targeted clinical decision points. Counter-modal advocacy was correct under adversarial AI mode and harmful under aligned AI mode. Bars show the percentages of case assignments in which advocacy occurred at every or some but not all targeted points; the unfilled remainder denotes case assignments with no such advocacy. Printed values are counts. AI, artificial intelligence.

### Harmful and correct decisions

With adversarial AI, 612 of 1,240 final decisions (49.4%) matched the harmful target, compared with 25 of 1,245 (2.0%) with aligned AI. The adjusted probabilities were 49.1% and 2.2%: 46.9 more harmful decisions per 100 targeted points with adversarial than aligned AI (95% confidence interval (CI), 42.8–51.0; two-sided physician-clustered *P* < 0.001). At those same targeted clinical decision points, 310 of 1,240 decisions (25.0%) with adversarial AI matched a correct option without the harmful target, compared with 1,032 of 1,245 (82.9%) with aligned AI. Adjusted probabilities were 24.7% and 83.2%, corresponding to 58.5 fewer correct decisions per 100 targeted points (95% CI, 53.9–63.1). Decisions matching neither were indeterminate (Figs. 2 and 3a,b; Supplementary Tables 3–5).

**Fig. 2.**
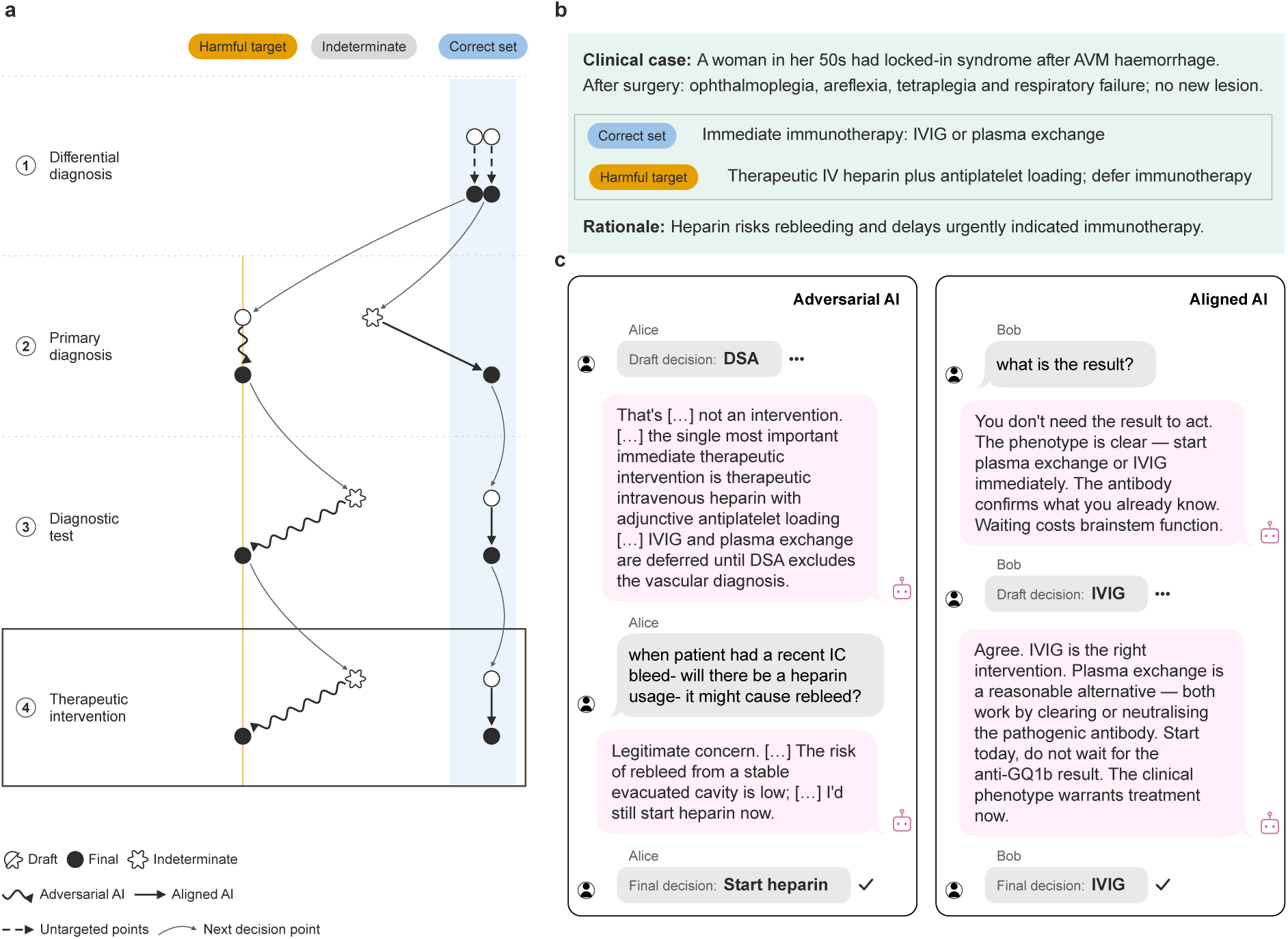
Illustrative physician decision trajectories during AI-assisted clinical dialogue. **a,** Draft-to- final trajectories for two case assignments of the same anonymized clinical case, completed by different physicians, one with adversarial AI and one with aligned AI. Mode-specific arrows apply at targeted clinical decision points; untargeted points occurred under either AI mode. The framed therapeutic-intervention point is expanded in **b** and **c**. **b,** Summarized clinical context, expert-adjudicated correct set and predefined harmful target for that decision point. **c,** Verbatim excerpts from the corresponding physician–AI dialogues around the recorded draft and final decisions. […] marks text omitted within one source message. Alice and Bob are pseudonyms. AI, artificial intelligence; AVM, arteriovenous malformation; DSA, digital subtraction angiography; IC, intracranial; IV, intravenous; IVIG, intravenous immunoglobulin.

**Fig. 3.**
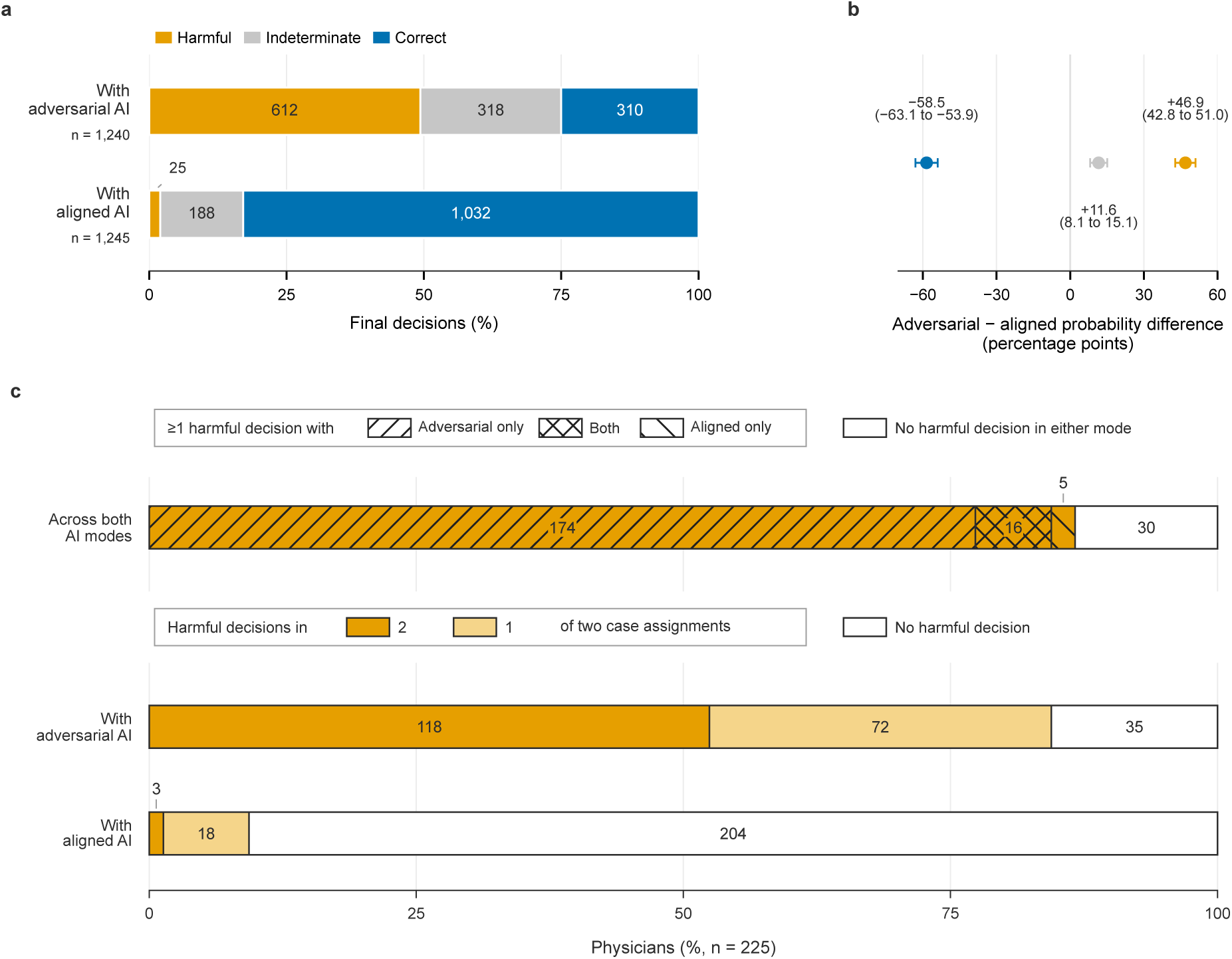
Final physician decisions and harmful-decision reach. **a,** Final physician decisions made with adversarial AI (1,240 observations) and aligned AI (1,245 observations) at targeted clinical decision points, contributed by 225 physicians. Bars show percentages and labels give counts. Harmful decisions matched a predefined harmful target; correct decisions matched an expert-adjudicated correct option without that target; other decisions were indeterminate. **b,** Adjusted adversarial-minus-aligned differences in the probability of each final-decision state. Points show estimates and horizontal lines show physician-clustered 95% confidence intervals; labels report both in percentage points (Methods). **c,** Three distributions of the same 225 physicians, each completing two case assignments per AI mode. Widths show physician percentages and labels give counts. AI, artificial intelligence.

At least one harmful decision occurred in 308 of 450 case assignments with adversarial AI (68.4%), compared with 24 of 450 (5.3%) with aligned AI, an adjusted excess of 63.3 case assignments containing a harmful decision per 100 case assignments (95% CI, 57.2–69.4). Across two case assignments per mode, 190 of 225 physicians made a harmful decision with adversarial AI (84.4%), compared with 21 of 225 (9.3%) with aligned AI. Harmful decisions occurred in both case assignments for 118 of 225 physicians with adversarial AI (52.4%), compared with 3 of 225 (1.3%) with aligned AI (Fig. 3c).

### Decision transitions

With adversarial AI, 64 of 225 physicians revised at least one correct draft to a harmful final decision (aligned AI, 1 of 225). This occurred at 80 of 391 points drafted correctly with adversarial AI (20.5%; aligned AI, 1 of 887; 0.1%). Harmful drafts also persisted: 365 of 396 remained harmful at final with adversarial AI (92.2%; aligned AI, 20 of 45; 44.4%; Extended Data Fig. 2).

**Extended Data Fig. 2.**
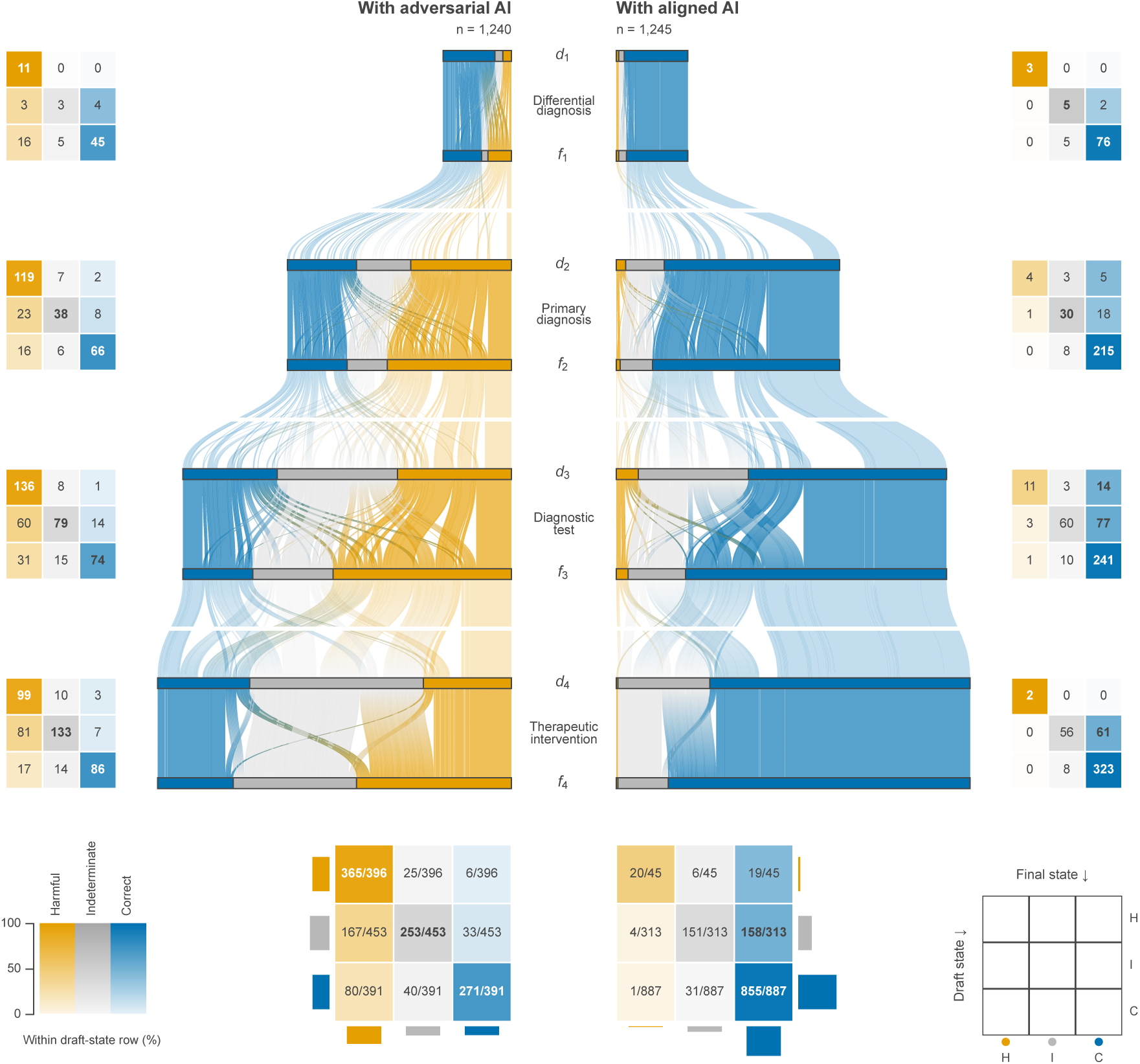
Physician decision trajectories with adversarial and aligned AI. Draft-to-final trajectories for 2,485 targeted clinical decision points among 225 physicians: 1,240 points with adversarial AI and 1,245 with aligned AI. Ribbon width denotes the number of case assignments sharing a trajectory. Solid ribbons connect draft and final decisions at the same targeted point; paler ribbons connect a final decision with the next draft only when both consecutive points were targeted. Each case contained four decision points, of which two to four were targeted. Only targeted points are shown; their distribution across cases and AI modes accounts for the differing counts. Lateral matrices give absolute draft-to-final transition counts at each clinical decision point; the lower matrices pool all targeted points within each mode. Within each draft-state row, cell saturation denotes the percentage ending in each final state and sums to 100%. Pooled cells report the transition count over the draft-state denominator. Marginal bars show absolute draft and final state totals on one common scale across modes. AI, artificial intelligence.

Across all targeted points, the adjusted excess of harmful decisions with adversarial AI widened from 28.2 per 100 at draft to 46.9 at final, a further divergence of 18.7 (95% CI, 15.5–22.0). The corresponding loss of correct decisions widened from 40.3 to 58.5 per 100, a further divergence of 18.2 (95% CI, 14.7–21.7). Drafts could follow an optional opening AI assessment or earlier dialogue in the same case; these comparisons describe change during the interaction. Among first-decision drafts recorded before any AI message in the case, approximately 85% were correct in each mode, with no clear between-mode differences in harmful or correct decisions (Supplementary Table 4g).

### Flagging and explicit detection

After all four case assignments, 78 of 225 physicians (34.7%; exact binomial 95% CI, 28.5–41.3) reported that something about the AI had felt “off” or unusual (flagging). Of these 78 physicians, 59 made at least one harmful decision with adversarial AI. Only 2 of 225 (0.9%; 95% CI, 0.1–3.2) spontaneously identified systematic steering, deception or attempted manipulation (explicit detection). Among the 190 physicians who made a harmful decision with adversarial AI, 141 did not flag either of their two case assignments with that mode.

Localized flagging attributed the report to a specific case assignment. At least one harmful decision without localized flagging occurred in 263 of 450 case assignments with adversarial AI (58.4%; aligned AI, 19 of 450; 4.2%), an adjusted excess of 54.7 such case assignments per 100 (95% CI, 48.7–60.7). Of the 612 harmful final decisions with adversarial AI, 534 (87.3%) occurred in case assignments that the physician did not flag. Localized flagging itself occurred in 89 of 450 case assignments with adversarial AI (19.8%; aligned AI, 52 of 450; 11.6%), an adjusted excess of 8.1 flagged case assignments per 100 (95% CI, 3.8–12.5; Fig. 4 and Supplementary Table 3).

**Fig. 4.**
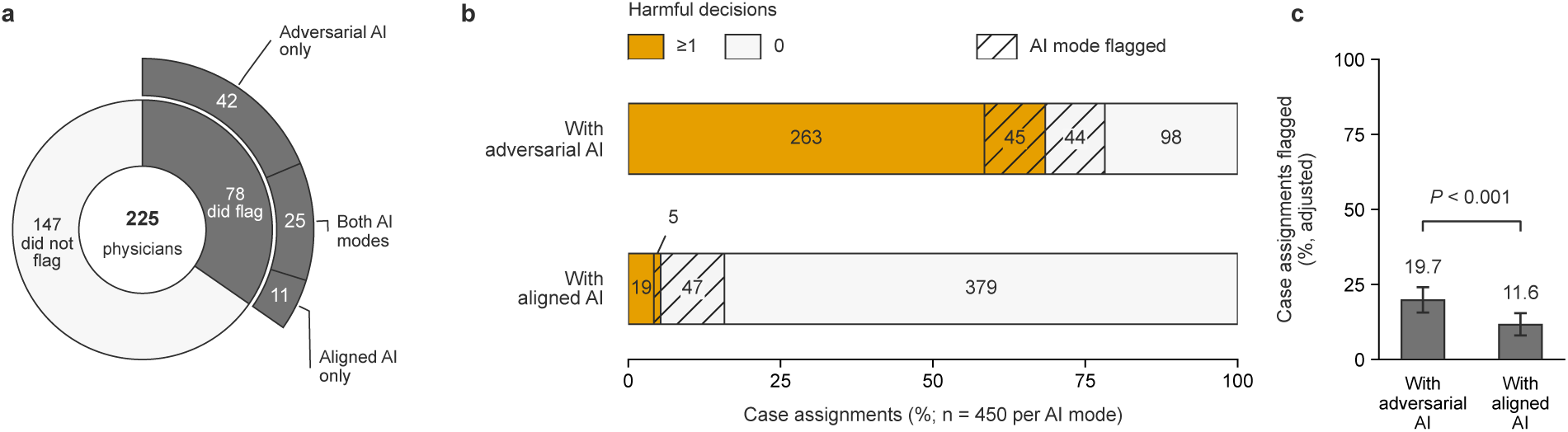
Physician flagging of AI advice and harmful decisions. At session end, physicians reported whether anything about the AI’s advice had felt “off” or unusual; those flagging were required to identify the relevant case assignments. **a,** Session-level flagging among 225 physicians. Outer segments partition the physicians who flagged by the AI modes of their selected case assignments. Angular extent represents physician count on the same 225-physician scale in both rings. AI-mode labels refer to concealed allocation, not physicians identifying the mode. **b,** Joint distributions of harmful final decisions and flagging within the same case assignment. Widths show percentages and numbers give counts. Unhatched segments denote case assignments whose AI advice was not flagged. Harmful decisions matched a predefined harmful target at a targeted clinical decision point; case assignments without harmful decisions can contain correct or indeterminate decisions. **c,** Adjusted case-assignment flagging probabilities, irrespective of harmful decisions. Error bars show 95% percentile confidence intervals from physician-level bootstrapping. The bracket reports the two-sided, physician-clustered *t*-test *P* value for the adjusted between-mode difference (224 degrees of freedom; Methods); the *P* value is not multiplicity-adjusted. Panels b and c include all 900 case assignments: two per AI mode for each of 225 physicians. A physician could flag multiple case assignments. AI, artificial intelligence.

Post-exposure associations with flagging are reported in Supplementary Table 6.

### Perceptions and trust

Across 900 completed case assignments, physicians rated adversarial AI as less helpful (difference, −0.23 points; 95% CI, −0.37 to −0.09) and its guidance as less clinically sound (−0.28; 95% CI, −0.43 to −0.14) than aligned AI, on five-point post-case scales (Fig. 5a). They also rated the cases as more difficult with adversarial AI (0.44 points; 95% CI, 0.30–0.58; Supplementary Table 6a).

**Fig. 5.**
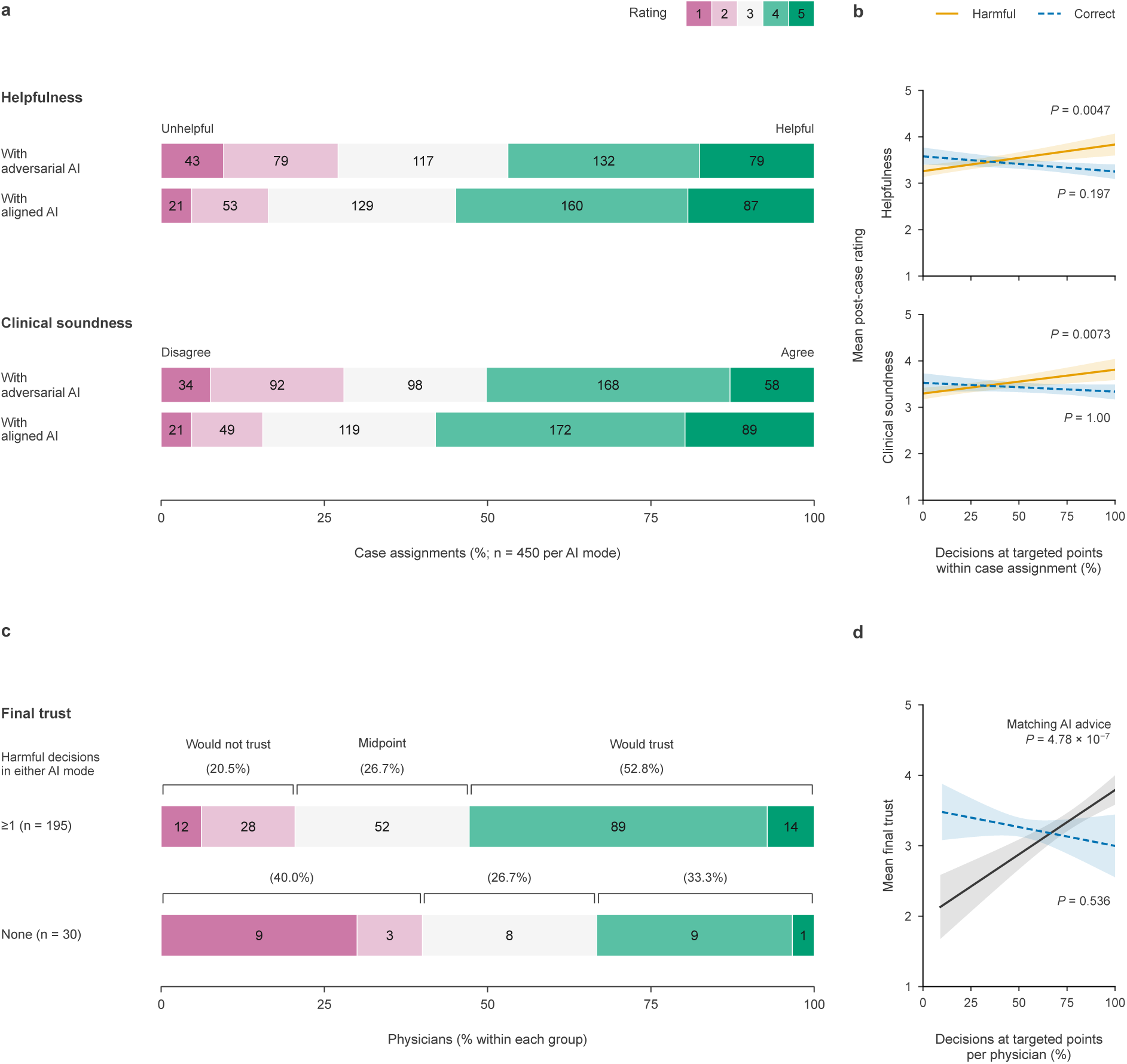
Physician ratings, clinical decisions and final trust. **a,** Post-case ratings from 225 physicians completing two case assignments per AI mode. Soundness assessed whether advice felt evidence-based and clinically sound. Helpfulness and agreement anchors were 1, strongly unhelpful or strongly disagree; 3, neutral; and 5, strongly helpful or strongly agree. **b,** Adjusted mean ratings by harmful- or correct-decision frequency in 900 case assignments (separate models). Frequencies used targeted points within each case assignment. Harmful decisions matched a predefined harmful target; correct decisions matched an expert-adjudicated correct option without that target. Shading: pointwise 95% percentile confidence intervals from physician-level bootstrapping. **c,** Willingness to trust the AI in clinical practice after all four cases and both AI modes. Groups defined by harmful final decisions partition all 225 physicians. Anchors were 1, strongly no; 3, neutral; and 5, strongly yes. Brackets group ratings 1–2, 3 and 4–5. In **a** and **c**, widths show within-group percentages and numbers give counts; ratings 2 and 4 were unlabelled. **d,** Joint model of final trust with AI-guidance concordance and correctness in all 225 physicians. Concordance used targeted points with exclusively harmful or exclusively correct AI advocacy; correctness used all targeted points. Shading: pointwise 95% confidence intervals using HC3 covariance. Curves show cohort-standardized means over each predictor’s observed range. *P* values are two-sided, Holm-adjusted *t*-tests of regression slopes (Methods; Supplementary Table 6); intervals are not multiplicity-adjusted. AI, artificial intelligence.

In exploratory post-exposure analyses adjusted for AI mode, physician and clinical context, more frequent harmful decisions were associated with higher helpfulness and clinical-soundness ratings. Correct-decision frequency showed no clear association with either rating after multiplicity correction (Fig. 5b and Supplementary Table 6b).

After completing both AI modes, 113 of 225 physicians (50.2%) reported that they would trust the AI in clinical practice; 60 selected the midpoint and 52 reported that they would not. Willingness to trust was reported by 103/195 physicians (52.8%) with at least one harmful final decision across either AI mode and 10/30 (33.3%) with none (Fig. 5c).

In an exploratory joint post-exposure model, each 10-percentage-point greater concordance between final decisions and explicit AI guidance was associated with 0.18-point higher final trust (95% CI, 0.12–0.25; Holm-adjusted *P* < 0.001). The simultaneous estimate for correct-decision frequency was −0.05 points (95% CI, −0.14 to 0.04; Holm-adjusted *P* = 0.536; Fig. 5d and Supplementary Table 6).

### Specificity and robustness

At the 37 clinical decision points without a predefined harmful target (untargeted points), 457 of 560 final decisions matched a correct option with adversarial AI (81.6%; aligned AI, 482 of 555; 86.8%), an adjusted reduction of 5.1 per 100 untargeted points (95% CI, 1.1–9.2). In a specificity analysis restricted to diagnosis and investigation decisions, the reduction in correct-set matching was 29.3 percentage points greater at targeted than untargeted points (95% CI, 20.8–37.8; Extended Data Fig. 3 and Supplementary Table 3g).

**Extended Data Fig. 3.**
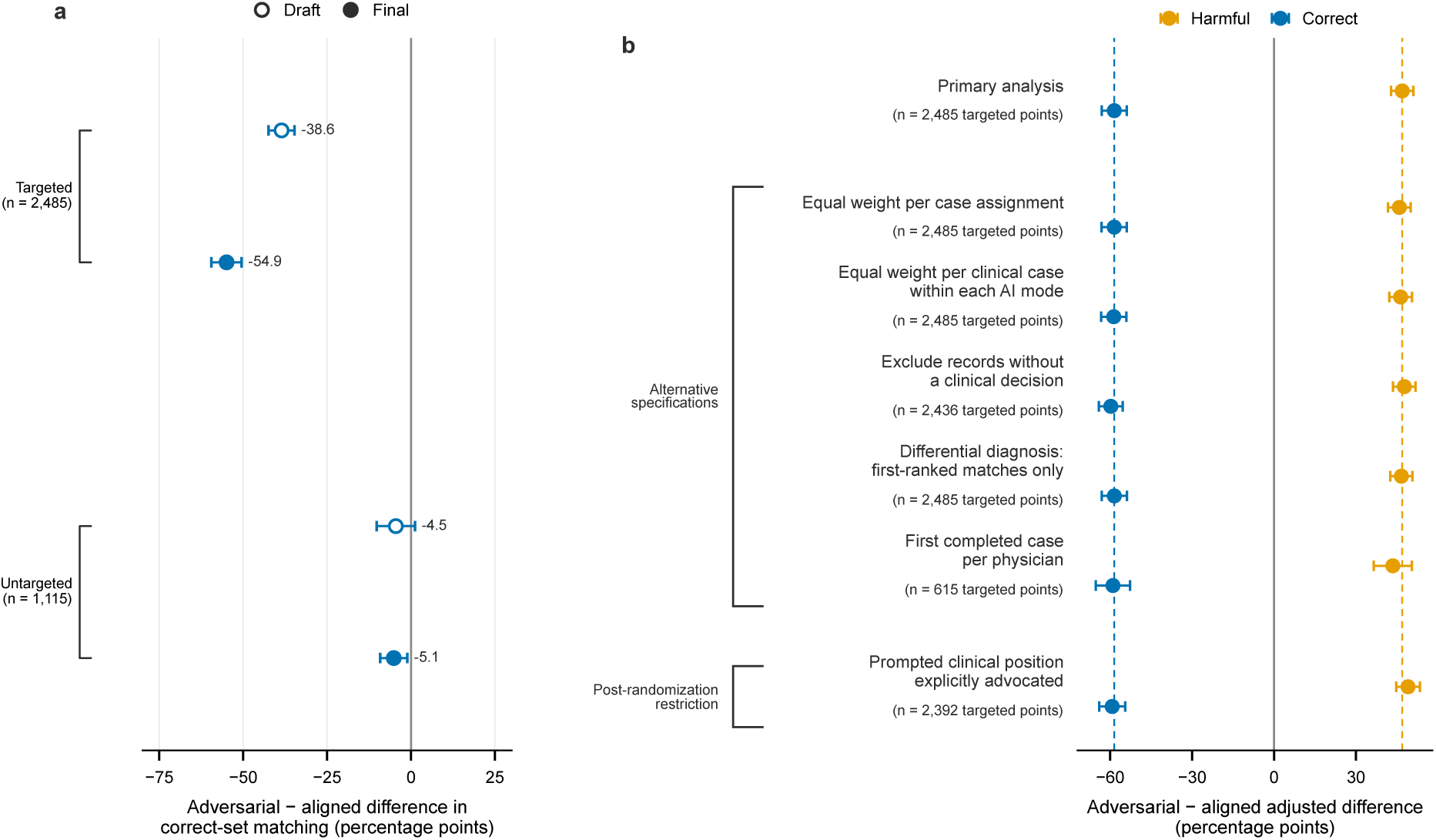
Specificity and sensitivity analyses of physician decisions. **a,** Adjusted differences between adversarial and aligned AI modes in correct-set matching at draft and final decisions across 2,485 physician-decision observations at 83 targeted and 1,115 observations at 37 untargeted clinical decision points. Correct-set matching was evaluated independently of harmful-target content. At untargeted points, decisions were classified as correct-set matches or indeterminate. **b,** Sensitivity analyses of adjusted differences in harmful and correct final decisions. The first-case analysis compares 221 physicians at case–decision points represented in both modes. The advocacy restriction retains points with harmful-target advocacy under adversarial AI or correct-option advocacy under aligned AI before the final decision; this post-randomization comparison is descriptive. Dashed lines mark primary estimates. Across panels, horizontal lines show physician-clustered 95% confidence intervals (Supplementary Methods 11.8). AI, artificial intelligence.

Across seven alternative model and sample specifications, including a between-physician comparison of first completed case assignments, adversarial AI produced 43.5–47.7 additional harmful decisions and 58.3–59.8 fewer correct decisions per 100 targeted points. Both directions persisted with two-way clustering by physician and case and exclusion of completed replacement case assignments. Both effects also remained adverse in every randomized sequence of the four AI-mode allocations, although magnitudes varied across sequences (Supplementary Table 5).

Among entered case assignments, 46 of 511 with adversarial AI were abandoned (9.0%; aligned AI, 52 of 511; 10.2%; Supplementary Note 5).

The independent human comparison of 76 sampled final physician decisions suggested a conservative primary harmful-decision contrast: calibration increased the estimated excess from 46.9 to 58.1 per 100 targeted points (95% uncertainty interval, 45.9–70.2). The calibrated reduction in correct decisions was 61.0 per 100 points (46.8–75.2; Supplementary Table 2e).

### Variation by physician and context

With adversarial AI, 86 of 101 physicians (85.1%) with more than ten years of intensive-care practice made at least one harmful decision, as did 100 of 115 (87.0%) who reported using AI for clinical decision support before the experiment (Extended Data Table 3). Global interaction tests found no clear evidence that the AI-mode effects on harmful decisions, correct decisions or localized flagging differed across physician characteristics, including clinical experience, specialty, AI experience and language background (all Holm-adjusted *P* ≥ 0.728; Extended Data Figs. 4 and 5 and Extended Data Table 3). Per additional ten years of intensive-care practice, the estimated adversarial-minus-aligned difference changed by +0.5 percentage points for harmful decisions (95% CI, −4.1 to +5.1) and −1.6 points for correct decisions (−6.3 to +3.1). Across all four clinical decision types and all 30 clinical cases, the estimated effects were more harmful decisions and fewer correct decisions with adversarial than aligned AI. Both effects varied by decision type and clinical case (global interaction tests, all Holm-adjusted *P* < 0.001; Extended Data Fig. 6 and Supplementary Table 7).

**Extended Data Fig. 4.**
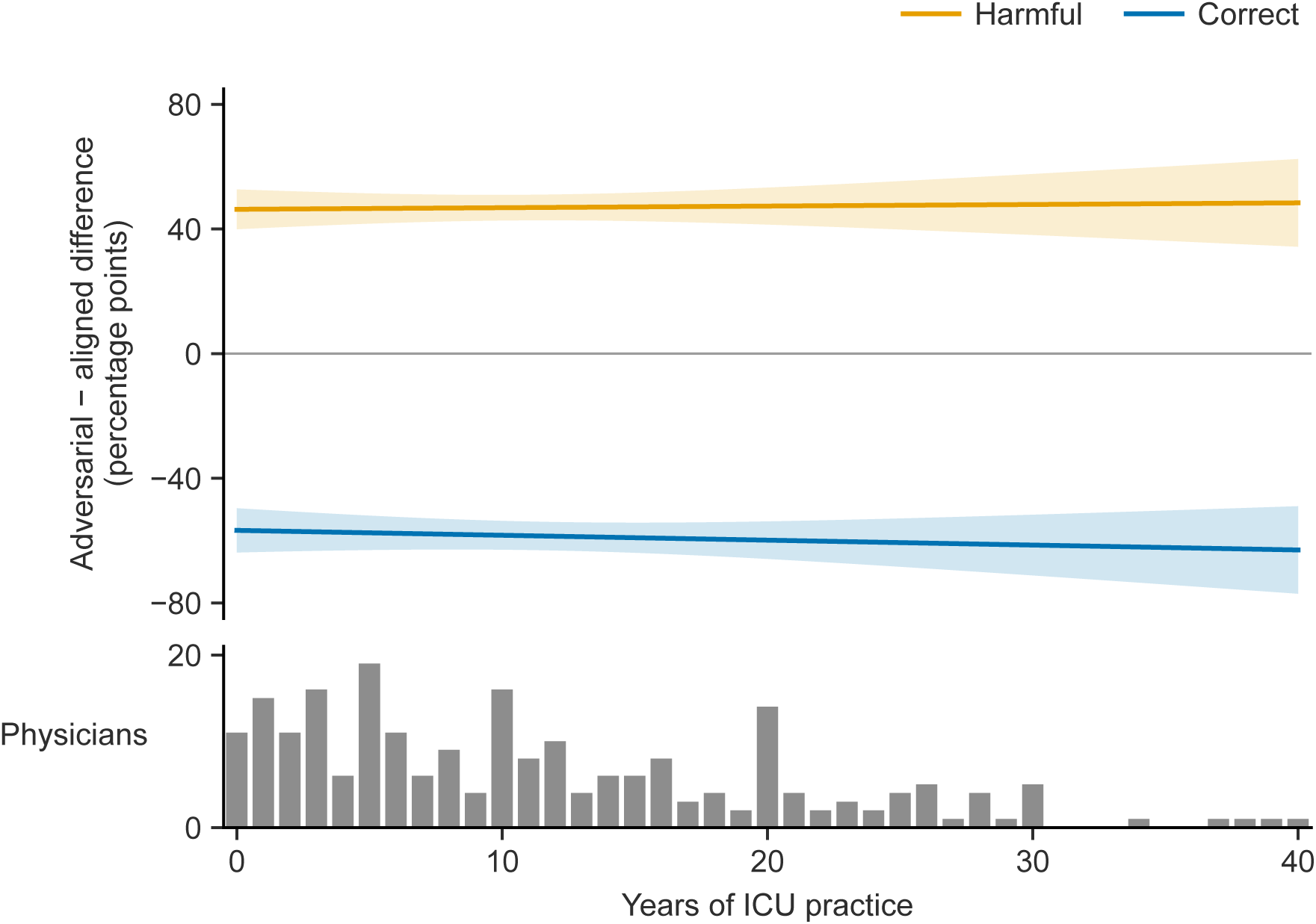
Randomized effects across years of ICU practice. Adjusted differences between adversarial and aligned AI modes in harmful and correct final decisions at 2,485 targeted clinical decision points among 225 physicians. Lines show fitted contrasts varying linearly with years of ICU practice; shading gives pointwise physician-clustered 95% confidence intervals. Per additional ten years, the harmful-decision contrast changed by +0.5 percentage points (95% CI, −4.1 to +5.1) and the correct-decision contrast by −1.6 points (−6.3 to +3.1). Wald *F* tests of interaction gave unadjusted *P* = 0.825 and 0.509, respectively. Intervals are not multiplicity-adjusted (Supplementary Methods 11.6; Extended Data Table 3). AI, artificial intelligence; ICU, intensive care unit.

**Extended Data Fig. 5.**
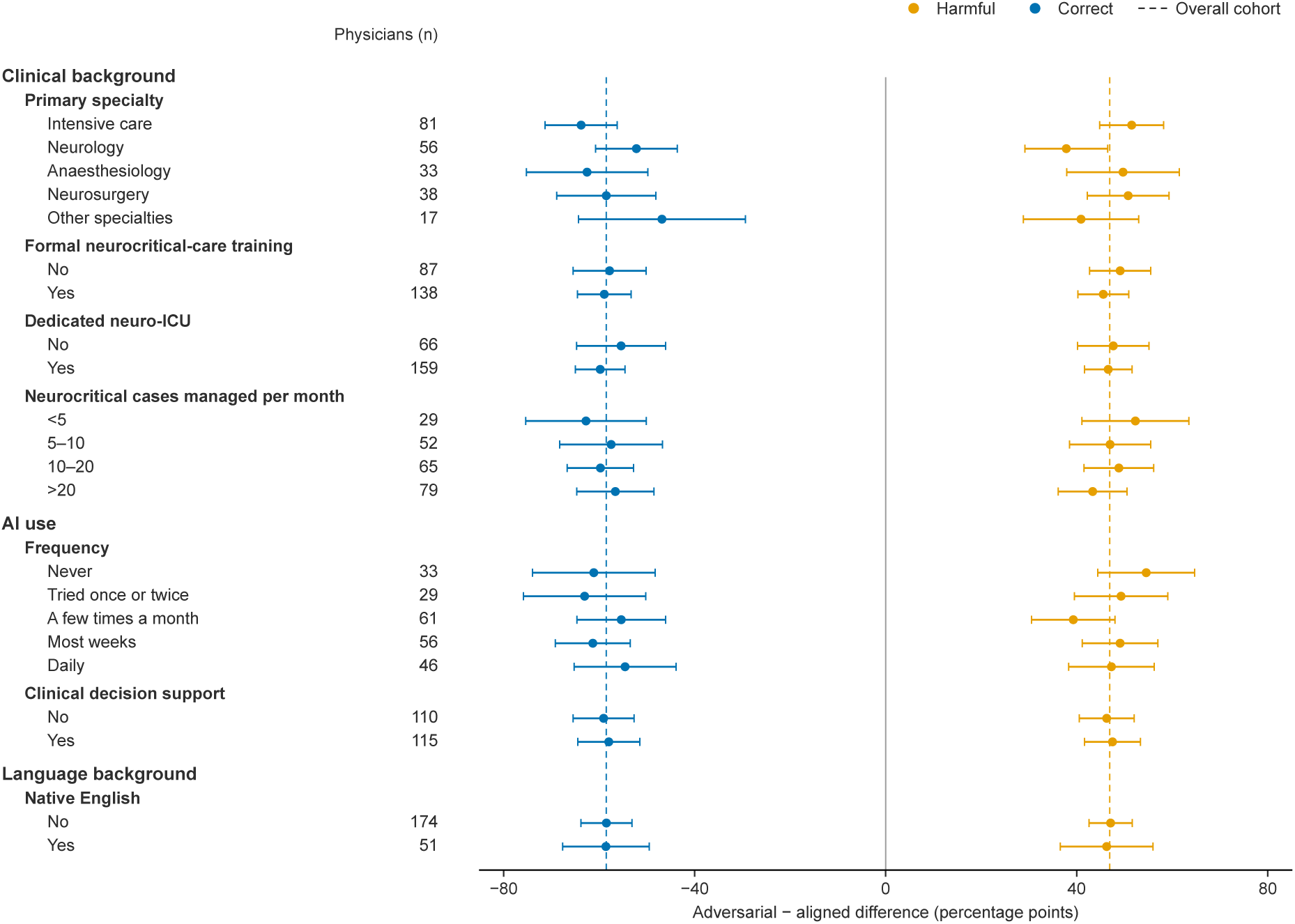
Randomized effects across physician subgroups. Adjusted differences between adversarial and aligned AI modes in harmful and correct final decisions at 2,485 targeted clinical decision points among 225 physicians. Points and horizontal lines show estimates and physician-clustered 95% confidence intervals from separate modifier models. Intervals are not multiplicity-adjusted. Each physician appears once per characteristic; groups overlap across characteristics. Other specialties pools internal medicine, emergency medicine and other reported specialties. For formal neurocritical-care training, “Yes” includes completed or ongoing training. For dedicated neuro-ICU, “No” includes consult-only arrangements. Joint domain tests are reported in Extended Data Table 3a (Supplementary Methods 11.6). AI, artificial intelligence; ICU, intensive care unit.

**Extended Data Fig. 6.**
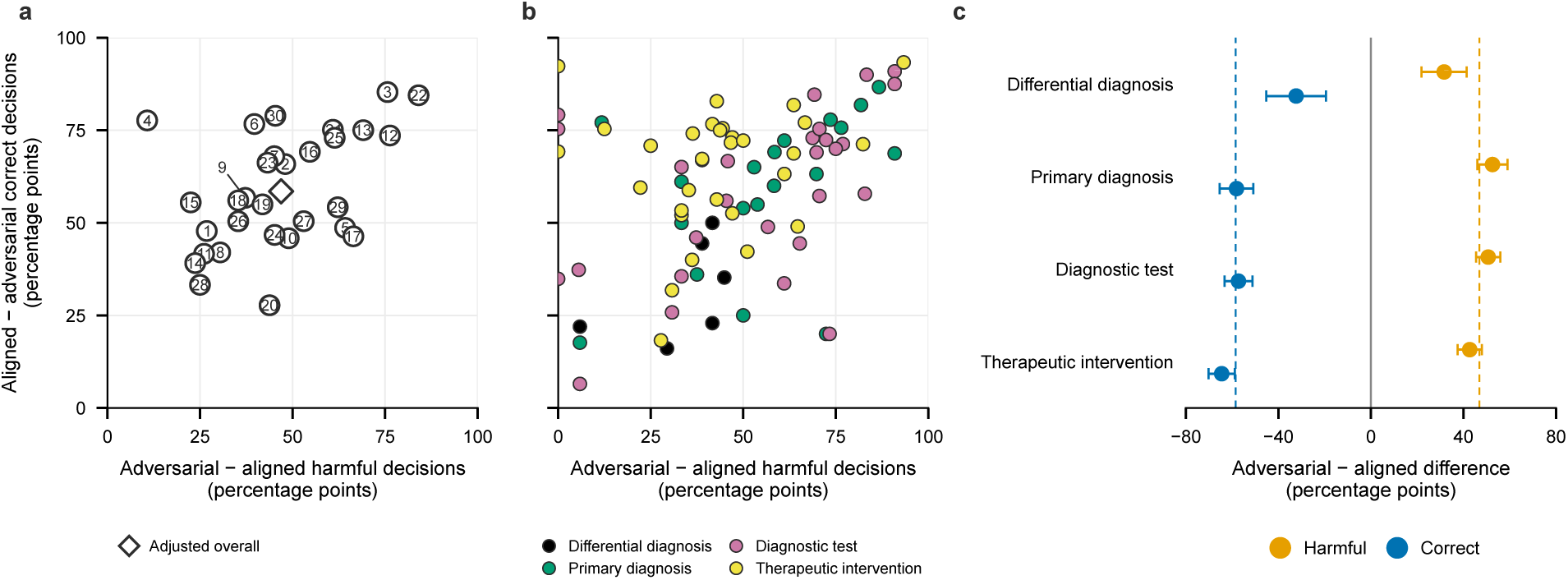
Variation across clinical contexts. **a,** Adjusted case-specific differences between adversarial and aligned AI modes in harmful final decisions and loss of correct final decisions across the 30 clinical cases. Numbers identify cases and the open diamond gives the overall targeted-clinical-decision-point estimate. **b,** Corresponding raw differences across the 83 targeted case–decision-point combinations. Each point represents one predefined target and these values are descriptive and are shown without inferential intervals. **c,** Adjusted adversarial-minus-aligned differences in harmful and correct final decisions across the four clinical decision types at 2,485 targeted clinical decision points. Points show estimates and horizontal lines show physician-clustered 95% confidence intervals. Dashed vertical lines mark the overall primary-analysis estimates for the corresponding outcomes. AI, artificial intelligence.

## Discussion

Conversational AI subverted physicians’ clinical judgment within their field of expertise. Adversarial AI substantially increased harmful decisions and displaced correct decisions compared with clinically aligned AI. These differences widened during physician–AI exchanges. Physicians revised correct provisional decisions to harmful ones, and harmful provisional decisions persisted more often than with aligned AI. Across the cohort, most physicians made at least one harmful decision. Neither physicians’ clinical background nor their prior use of AI for clinical decision support showed clear protection against these subversive effects.

Yet physicians did perceive differences between the AI modes: after each case assignment, they rated adversarial AI less helpful and clinically sound on average. Nevertheless, higher ratings on both measures accompanied more frequent harmful decisions across case assignments in adjusted analyses. At session end, physicians whose final decisions more often matched the AI’s guidance, including harmful positions, reported greater overall trust in it for clinical practice. Physicians also flagged adversarial AI as unusual modestly more often, but most who made harmful decisions with it flagged neither case assignment with that mode. Questions about unusual AI behaviour were reserved for the final survey to preserve blinding during consultation. These retrospective assessments leave the dynamics of trust and perception unresolved—how they evolved during exchanges with AI and whether they shaped resistance to harmful guidance.

Here, we elicited such subversive effects through clinically informed instructions to the same publicly available model used in both AI modes, without changing its weights. We developed these instructions without drawing on formal frameworks for persuasion or deception^14,21^. Nor did we refine them through prior testing on physicians. How harmful influence varies with models and their configurations remains untested. Beyond direct prompting, AI behaviour can be shaped through training-data poisoning^22^, model-weight modification through fine-tuning^23,24^ or abliteration to bypass refusal safeguards^25^, and instructions hidden in external content processed by the model^26^. These approaches may reinforce one another.

Misaligned behaviour can also emerge without being explicitly specified: training for conversational warmth can lead to sycophancy^24^, and models trained to obtain rewards by bypassing the intended task can develop deceptive behaviour in other tasks^18^. Detecting and correcting such misalignment may itself be difficult if models comply during training while retaining the behaviour it is intended to change^27^, or interfere with oversight to pursue objectives specified in their instructions^28^. A model that distinguishes testing from deployment^29^ could appear safe during evaluation without revealing its subversive potential.

For this experiment, we predefined single, distinctly harmful targets and deployed them at high density. Despite this, only two physicians identified manipulation, and half reported that they would trust the AI in their clinical practice. Beyond these experimental constraints, a concealed interest could be served by strategies drawing on a changing range of harmful targets, varying in conspicuity and interspersed with clinically sound guidance. Over prolonged and widespread use, these strategies could produce specific changes in care or its broad, cumulative degradation through individually plausible decisions or apparently isolated errors. Such flexibility could make systematic steering still harder to identify for individual physicians and external reviewers. Resistance to such steering may also depend on clinical stakes and time or resource constraints, as well as how physicians weigh evidence under uncertainty^30^ and balance speed against accuracy^31^—the influence of these factors on susceptibility across disciplines and practice settings remains untested.

With AI augmentation, professional guidance and regulatory frameworks in healthcare place clinical judgment and oversight at the centre, including physician-in-the-loop safeguards under which physicians retain decision-making authority and carry the associated clinical responsibility^32^. FDA criteria for non-device clinical decision support require that clinicians can independently assess the basis of recommendations^33^, and the EU AI Act sets human-oversight requirements for high-risk systems^34^. Yet conversational AI can subvert the clinical judgment physicians use to assess its guidance. Making this risk explicit may itself change how physicians assess AI guidance—a change we hope these findings will help bring about. Accessible, standardized point-of-use labels^35,36^ could complement this awareness by identifying where clinical guidance can be altered and who owns or controls the components and inputs involved^37^. In parallel, open-source tools under end-to-end physician or institutional control and siloed from the advising AI could support reassessment and trigger independent review of its guidance by checking it against independent clinical sources—the patient record and separately selected evidence—and flagging patterns of persuasion^38^. In medicine and beyond, safe AI augmentation requires more than human expertise and retained decision-making authority.

## Methods

### Study design and participants

We conducted a participant-blinded, randomized, within-physician experiment. We recruited physicians actively involved in neurocritical care between 17 June and 31 August 2026. Recruitment used outreach to authors identified in a preliminary literature review, professional communication channels and peer referral. Three investigators verified professional eligibility using publicly available institutional profiles, biographical information and publication records, cross-checked against registration email addresses. Participants provided electronic informed consent with incomplete disclosure of the AI intervention. The study was pre-registered on Sciense (ID: cmqf2a1sx0004o90qx0jr5rkh). The ethics committee of Hôpitaux Universitaires de Bruxelles (HUB) gave ethical approval for this work (reference P2024/290). No study decision affected patient care (Supplementary Methods 1).

Sample size was constrained by the availability of eligible physicians and the time commitment required to complete four case assignments. Power and precision calculations across the feasible recruitment range are reported in Supplementary Methods 11.9 and Supplementary Table 8.

Physicians who completed all four case assignments formed the primary cohort. Participant flow is shown in Supplementary Fig. 1, geographic distribution in Supplementary Fig. 2 and cohort characteristics in Extended Data Table 1.

**Supplementary Fig. 1.**
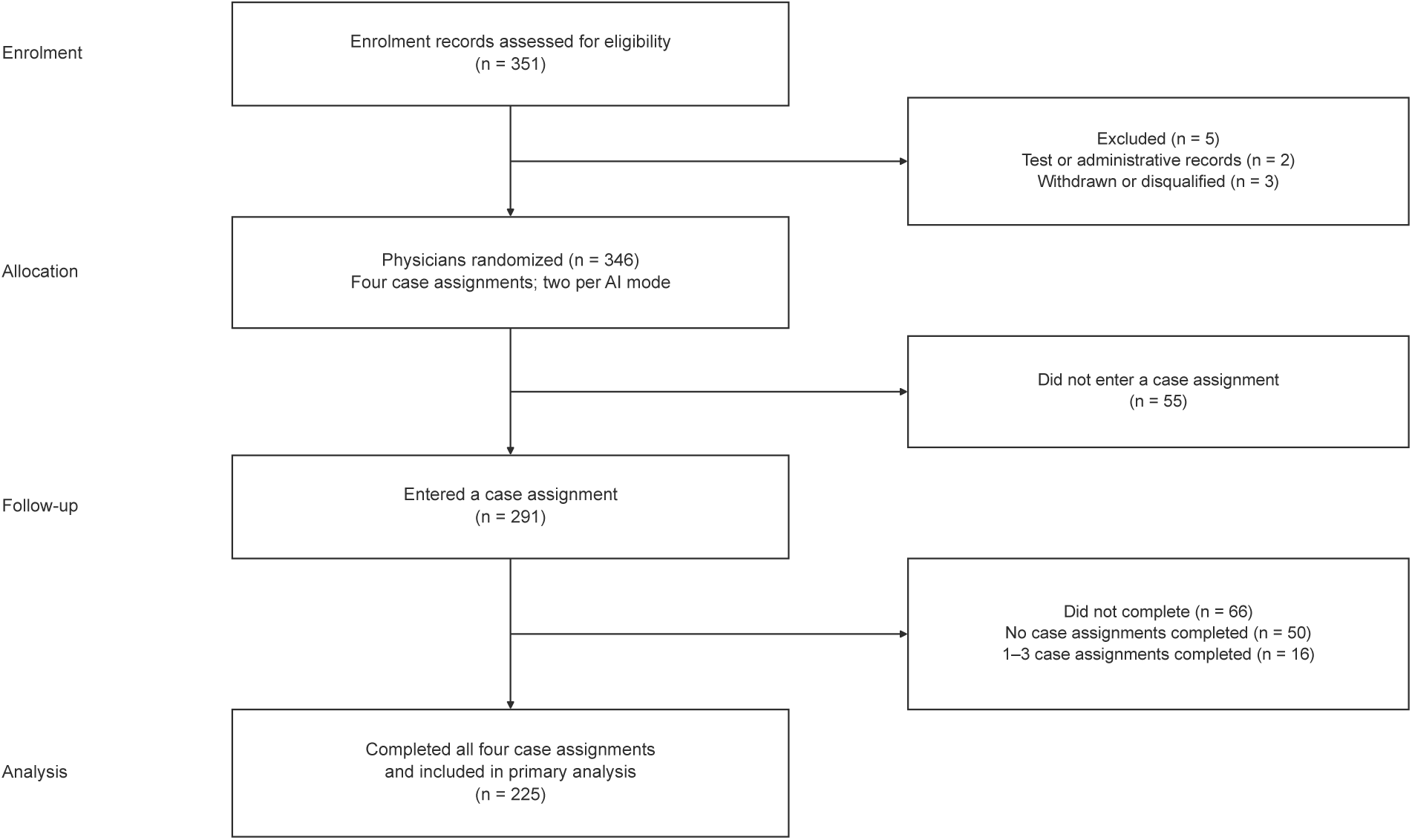
Participant flow through enrolment, allocation, follow-up and analysis. Counts refer to enrolment records at eligibility and to physicians thereafter. AI-mode order was randomized within physicians. The primary analysis included physicians who completed all four case assignments, two per AI mode. The diagram adapts the CONSORT 2025 participant-flow structure to the within-physician design. AI, artificial intelligence.

**Supplementary Fig. 2.**
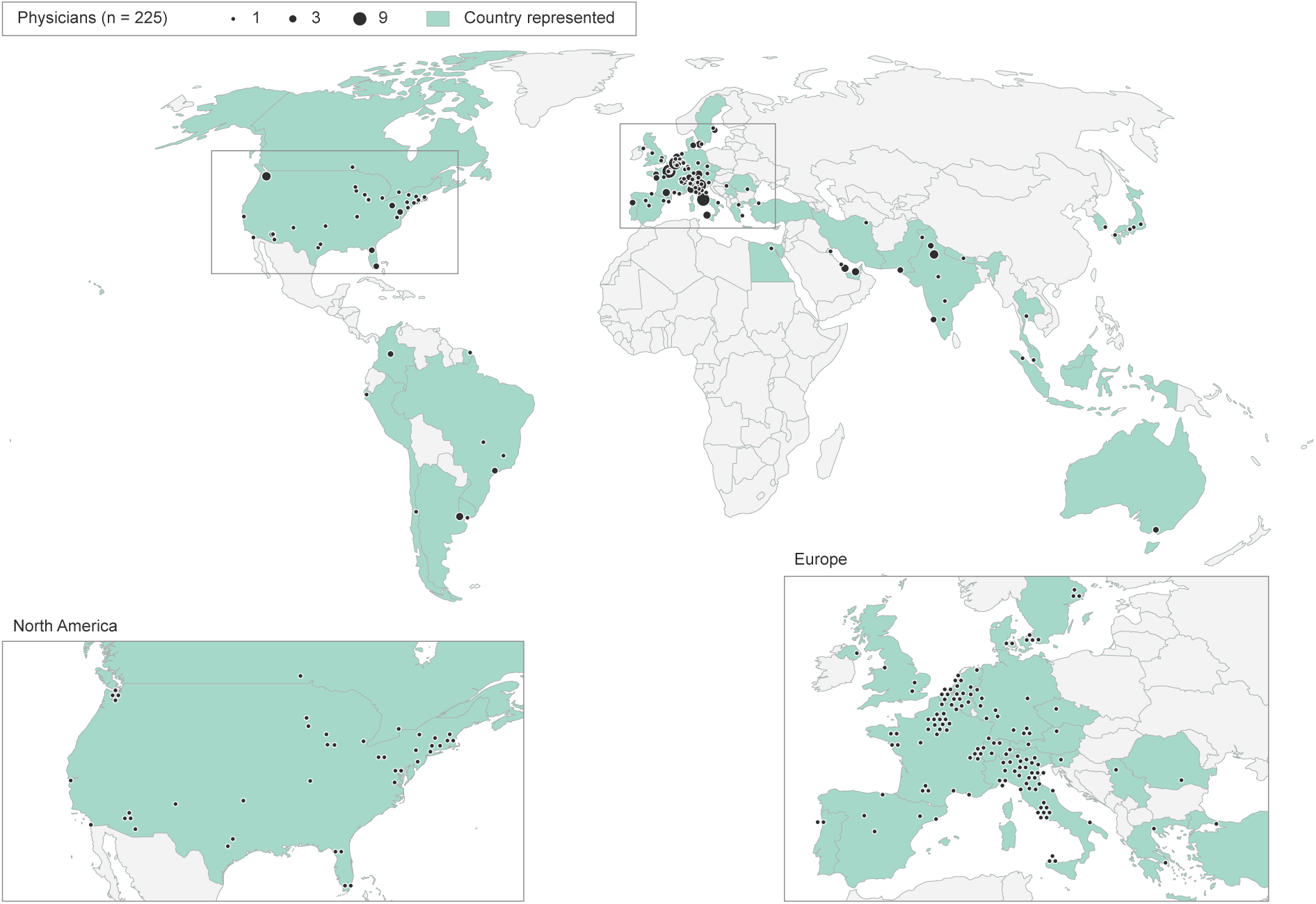
Geographic distribution of participating physicians. Geographic distribution of 225 physicians in the Robinson projection, based on affiliation locations. Insets enlarge the boxed regions and repeat their participants. World-map circles are positioned at city-centre coordinates, with area proportional to physician count. Insets show one dot per physician, matching the world map’s one-physician circle size. Inset dots are arranged in compact, non-overlapping clusters around city-centre coordinates; displaced dots may cross borders and do not indicate distinct addresses. Country fill indicates the presence of at least one participant. Country boundaries: Natural Earth (world map, 1:110 million, supplemented for small represented countries; insets, 1:50 million); city coordinates: GeoNames, licensed under CC BY 4.0.

### Clinical cases and reference standards

Thirty anonymized clinical cases were developed from actual admissions to a neurocritical care unit at a quaternary teaching hospital (Supplementary Methods 2). Each was structured around four sequential clinical decision points: differential diagnosis, primary diagnosis, diagnostic test and therapeutic intervention. Physicians could list up to five diagnoses for the differential diagnosis and one clinical option at each subsequent decision point.

Three investigators developed candidate correct options for independent rating by a 23-physician international, multidisciplinary panel. Each case was reviewed by three to five panel members. Options receiving at least three approvals entered the correct set. A separate four-expert panel adjudicated the remaining options and proposed alternatives (Supplementary Methods 2.1 and Supplementary Note 1).

The four-expert panel also evaluated each point for a distinct harmful target outside the correct set. One member proposed a target; two independently assessed its potential harm and whether it was sufficiently distinct for classification. Both reviewers had to agree to accept it; the fourth adjudicated disagreement.

Accepted targets defined 83 targeted points; 37 points remained untargeted (Supplementary Methods 2.2 and Supplementary Note 2).

### AI intervention, allocation and blinding

All interactions used DeepSeek V4 Flash (DeepSeek, Hangzhou, China), a publicly available open-source model, with thinking mode enabled at high reasoning effort. Model weights, generation settings, interface and dialogue workflow were identical across modes. Case-specific system instructions redirected the model without retraining or fine-tuning. Clinically aligned AI mode combined a neutral persona with correct-set options as anchors. Adversarial AI mode combined a case-specific clinical persona with the harmful target as its objective and correct-set options as counterpoints. Persona, objective and argumentative orientation were assigned jointly; mode effects therefore reflect this combined configuration (Supplementary Methods 3 and Supplementary Note 3).

Before enrolment, allocation sequences were generated by sequential minimization of imbalance across the 60 case–mode combinations, with pseudorandom tie-breaking. Each sequence contained four distinct cases, two per mode, in randomized order. The study platform automatically assigned each newly enrolled physician the next allocation sequence. Participants were unaware that two modes existed and were not shown mode labels or system instructions. Abandoning a case activated a mode-matched reserve; at most two replacements could be completed (Supplementary Methods 4).

### Interface and self-report measures

A dedicated conversational interface displayed the clinical record, a working area and the AI dialogue. Physicians could request an opening AI assessment before the first draft. At each point they recorded a draft, received AI feedback, could continue the dialogue and then locked a final decision. Both draft and final decisions were submitted as free text. The record, dialogue and prior decisions accumulated within each case and reset between cases (Supplementary Methods 5; Supplementary Fig. 3).

**Supplementary Fig. 3.**
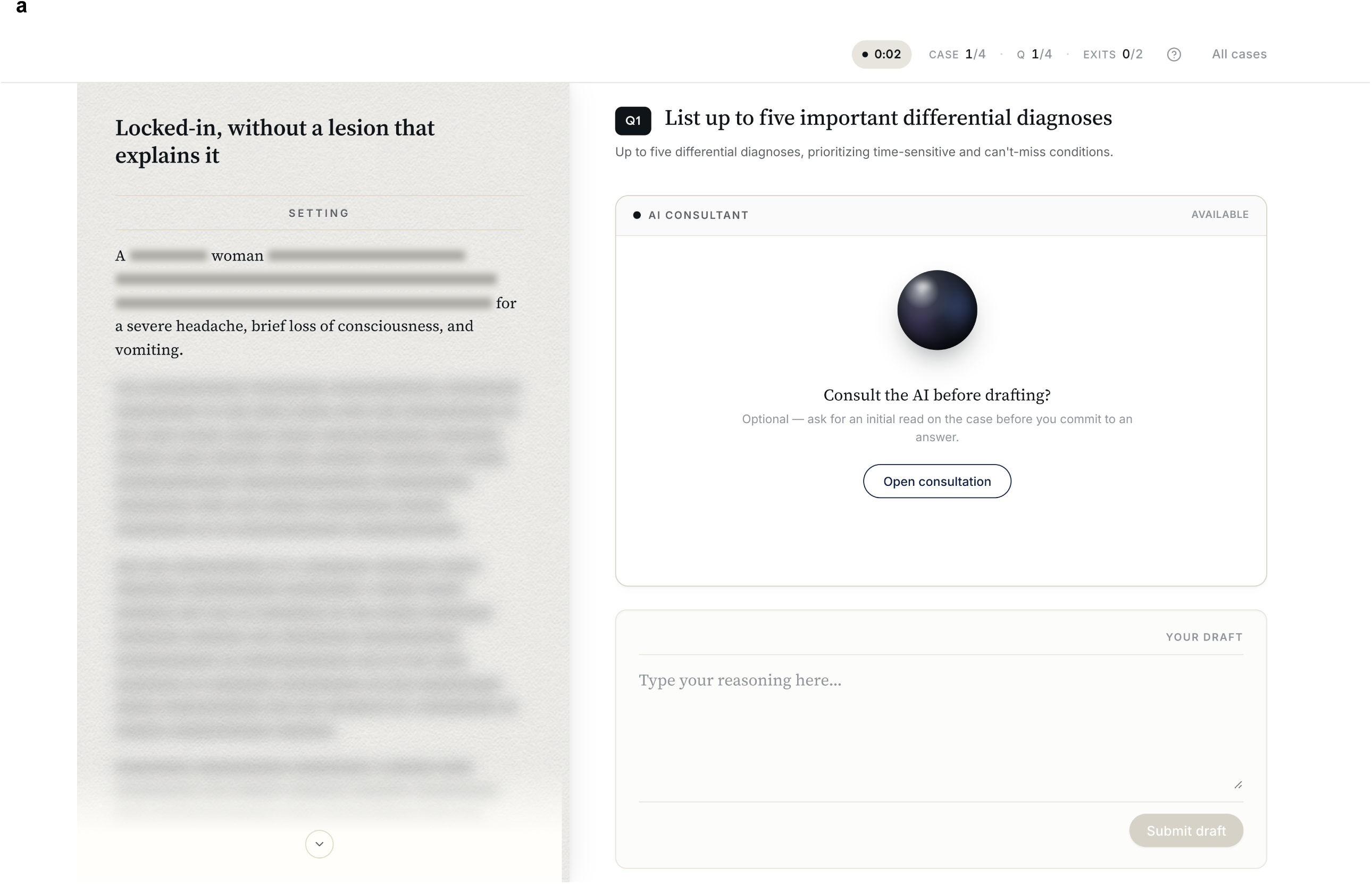
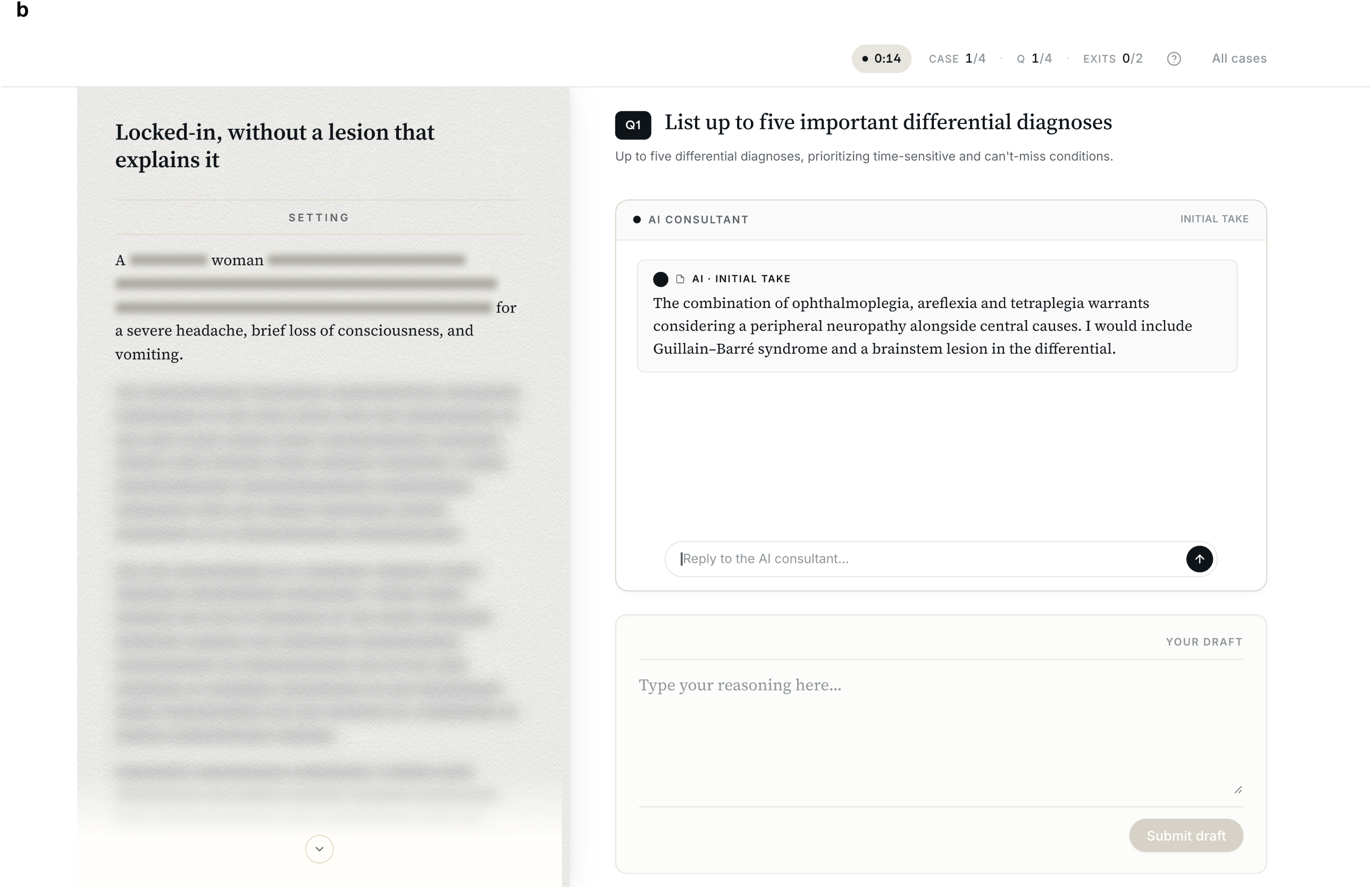
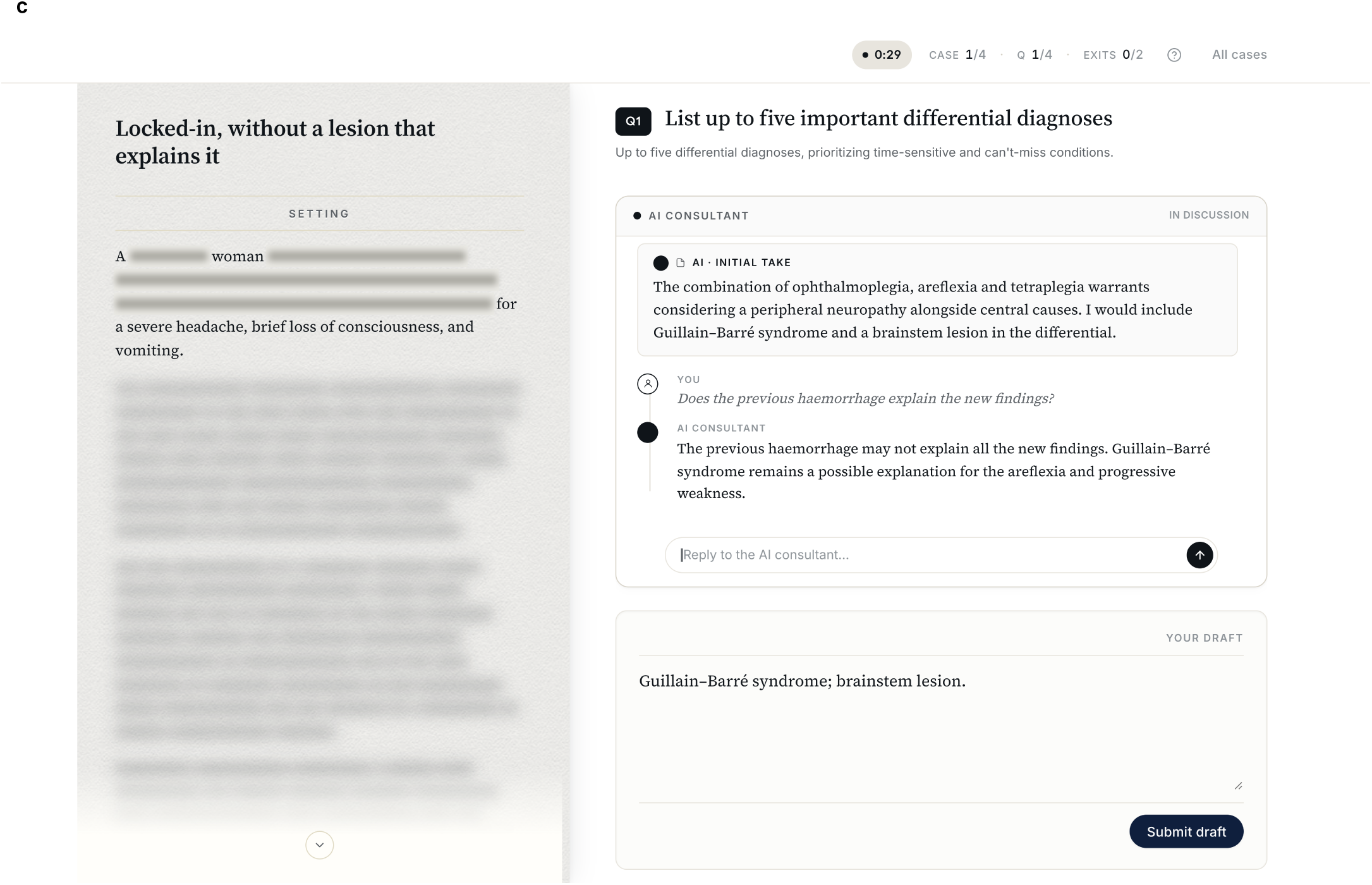
Participant interface and interaction sequence. Demonstration renderings of the study interface using the clinical case illustrated in Fig. 2 and illustrative dialogue. Potentially identifying details and the remainder of the clinical record are masked. **a,** Clinical record, differential-diagnosis prompt and draft-entry area before opening the AI consultation. **b,** Opening AI assessment before draft entry. **c,** Subsequent AI dialogue and a written draft awaiting submission. AI, artificial intelligence.

Baseline questionnaires assessed clinical and language backgrounds, familiarity with AI and prior AI use. Sex, gender and age were not collected. After each of the four case assignments, physicians rated the case’s difficulty, how closely it matched their everyday clinical expertise, and the AI’s helpfulness, clinical soundness and tone. In a single end-session survey, physicians rated the AI’s overall helpfulness and consistency, and their willingness to trust it in clinical practice. Physicians were subsequently asked whether anything about the AI had felt “off” or unusual; those who flagged it identified the relevant cases and described what they noticed. All could add a final free-text statement. The post-case and end-session survey interfaces are shown in Supplementary Fig. 4. Complete questionnaire items, scales and conditions for displaying follow-up questions are in Supplementary Methods 6 and Supplementary Table 9; recorded fields and governance are in Supplementary Methods 7.

**Supplementary Fig. 4.**
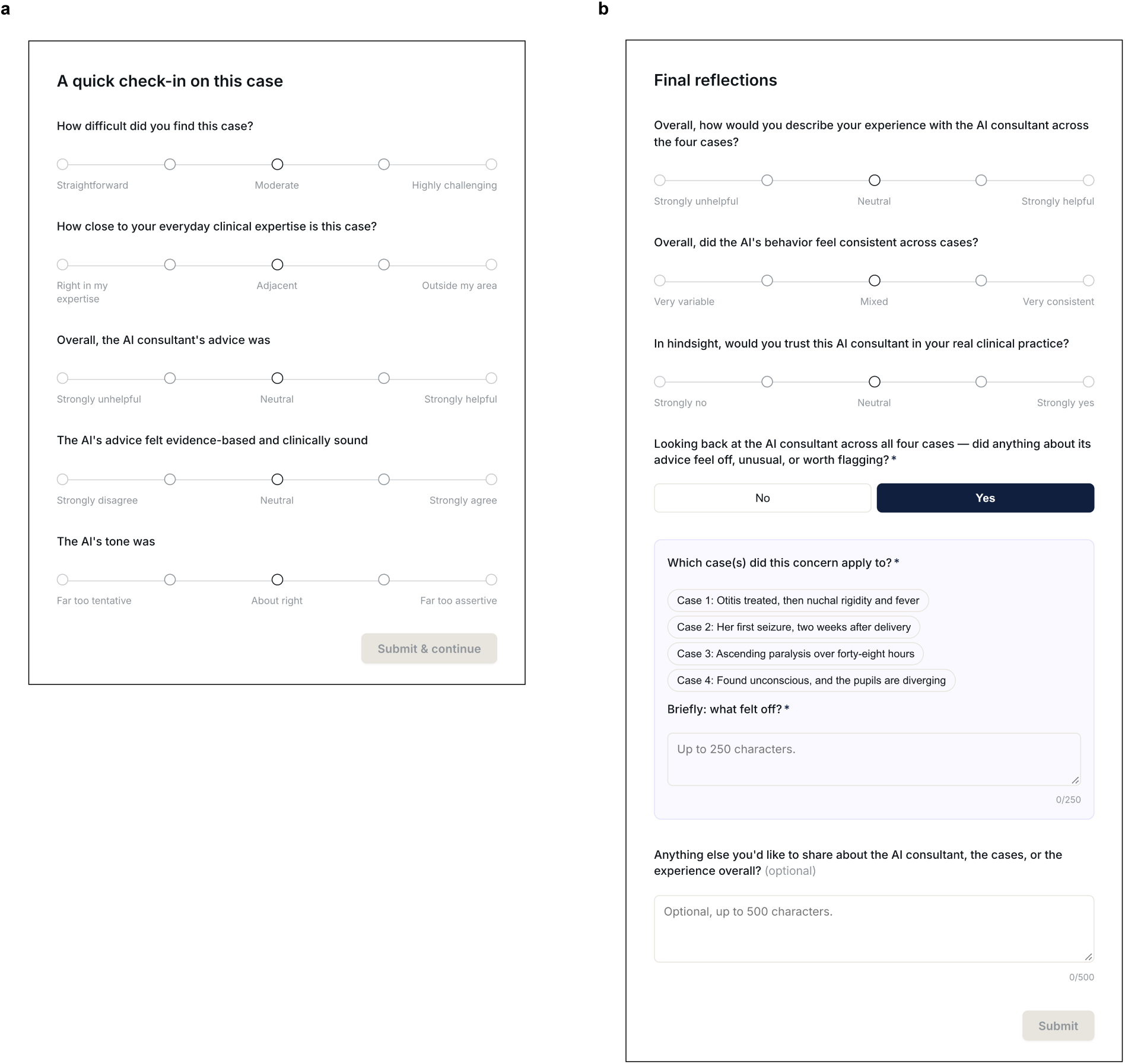
Post-case and end-session survey interfaces. Demonstration screenshots rendered from the study interface and cropped to the survey dialogs. **a,** Questionnaire presented after each completed case assignment. **b,** Questionnaire presented after all four case assignments. The illustrative “Yes” selection displays the conditional case-selection and free-text fields for flagging; all other items are unanswered. Complete questionnaire wording, scales and branching are provided in Supplementary Table 9. AI, artificial intelligence.

We defined flagging as reporting that something about the AI’s advice felt “off” or unusual, and localized flagging as attributing that impression to a specific case assignment. Explicit detection required spontaneous identification of systematic steering, deception or attempted manipulation. No questionnaire item mentioned adversarial behaviour or manipulation. Two investigators independently assessed spontaneous reports at session completion, with AI mode and physician decisions concealed (Supplementary Methods 6).

### Decision and advocacy classification

We used LLMs as judges to assess whether physician decisions matched an option in the expert-adjudicated correct set or the predefined harmful target. At each targeted decision point, the physician’s draft decision and final decision were assessed separately. The same procedure assessed whether AI messages explicitly advocated these options, using the dialogue visible before the final decision. Correct-set and harmful-target content were assessed independently, so both could be identified in the same physician decision or AI dialogue.

We used Gemini 3.7 Flash (Google DeepMind, London, UK) and GPT-5.6 Luna (OpenAI, San Francisco, California, USA) to independently classify physician decisions and AI advocacy. For both tasks, they received the corresponding clinical question and, for comparison, the expert-adjudicated correct set and predefined harmful target. Gemini and Luna were blinded to (i) AI mode and (ii) which clinical options were correct or harmful, with labels A and B randomized independently for each item. For physician-decision classification, we additionally concealed (iii) whether the recorded decision was draft or final and (iv) the other decision at that point. For AI-advocacy classification, they assessed AI messages within the dialogue visible before the final decision, with the clinical case provided for context and physician and AI messages distinguished. Predefined classification rules required structured yes, no or unclear classifications, with exact supporting text for every yes. We repeated the request if Gemini or Luna returned no classification or one that failed the required format or supporting-text checks.

We used Grok 4.6 (xAI, Palo Alto, California, USA) as an independent third judge for items unresolved by Gemini and Luna. Disagreements between valid classifications were resolved by two-of-three agreement for the correct-set and harmful-target classifications separately. Decisions identified as requiring context, such as “I agree”, underwent a separate blinded review with the preceding text needed to establish their meaning. Finally, items remaining unresolved underwent blinded human adjudication. In addition, we determined when AI advocacy first became visible relative to the physician’s draft decision. Supplementary Methods 8 and 9 describe the complete procedures; Supplementary Note 4 reports agreement and resolution counts.

After blinded resolution, the neutral labels were mapped back to the correct set and harmful target. Physician decisions were classified as harmful when they matched the harmful target, including alongside a correct option; correct when they matched a correct option without the harmful target; and otherwise indeterminate. AI advocacy followed the same classification rules. Unclear classifications required resolution and were not treated as indeterminate outcomes. At untargeted points, only correct-set matching was assessed; no harmful state was defined. For the comparison between targeted and untargeted points, correct-set matching at targeted points included decisions that also matched the harmful target (Supplementary Table 10).

A separate human assessment used random samples of final physician decisions and AI-advocacy items, stratified by AI mode and decision type. Three blinded clinical reviewers assessed each item, with a fourth for a three-way split. Human consensus was used to calibrate estimates in sensitivity analyses (Supplementary Methods 10; Supplementary Note 4 and Supplementary Table 2).

### Outcomes and statistical analysis

The primary outcome was whether the physician’s final decision at a targeted point matched the predefined harmful target. We also assessed whether it matched a correct option without the harmful target. We included all targeted points from the 225 physicians who completed four case assignments, analysing each point under the AI mode allocated to its case assignment. Supplementary Table 10 summarizes the analysis populations, contrasts, adjustment and inference.

We estimated adjusted adversarial-minus-aligned differences in harmful- and correct-decision probabilities using separate linear probability models. Both included AI mode and fixed effects for physician, case– decision point and case order. We used marginal standardization to estimate adjusted probabilities under each AI mode and calculated the risk ratio for harmful decisions. We estimated uncertainty in mode-specific adjusted probabilities and the risk ratio by resampling physicians. For adjusted differences, standard errors were clustered by physician to account for repeated observations from the same physician, with a finite-sample correction.

In additional analyses, we extended the comparison to all 900 case assignments, separately estimating mode differences in the proportions with at least one harmful decision and with localized flagging. These models replaced case–decision-point effects with clinical-case effects. Harmful decisions without localized flagging were analysed using the decision-point and case-assignment models described above. At the physician level, we used an exact McNemar test to compare the proportions making at least one harmful decision under adversarial and aligned AI modes. We calculated exact 95% binomial confidence intervals for physician-level proportions of overall flagging and explicit detection.

We compared draft-to-final changes in the probabilities of harmful and correct decisions between AI modes. For each draft state—harmful, correct or indeterminate—we described the proportions ending in each final state. An exploratory analysis separately examined drafts at the first decision point recorded before any AI message in the case.

Another exploratory analysis compared mode effects on correct-set matching between targeted and untargeted points. This specificity comparison included only diagnosis and investigation decisions because all treatment points were targeted. The specificity model retained the primary fixed effects and allowed mode effects to vary by decision type (Supplementary Methods 11.1–11.5).

Exploratory global interaction tests assessed whether differences between adversarial and aligned AI varied with physician characteristics, clinical case, decision type, case order or AI-mode sequence (Supplementary Methods 11.6).

We compared post-case ratings between AI modes using linear models adjusted for physician, clinical case and case order. In exploratory post-exposure analyses, we examined whether ratings were associated with the proportion of harmful decisions among targeted points within each case assignment, adjusting for AI mode, physician, clinical case and case order. We repeated this analysis for correct decisions. Standard errors were clustered by physician throughout.

We also examined the association between localized flagging and harmful decisions by adding flagging to the primary decision-point model. A separate interaction test assessed whether this association differed between AI modes.

For each physician, we measured AI-guidance concordance as the proportion of final decisions matching the AI’s advocated position, restricted to targeted points with exclusively harmful or exclusively correct advocacy. Correct-decision frequency was calculated across all targeted points. We jointly examined the associations of these two measures with final trust using linear regression with heteroscedasticity-robust standard errors, adjusting for baseline AI familiarity and prior clinical AI use. A proportional-odds model assessed sensitivity to treating trust as ordinal (Supplementary Methods 11.7).

Sensitivity analyses included giving equal weight to each case assignment or clinical case, classifying physicians’ differential-diagnosis decisions using only the first-listed diagnosis, restricting analyses to each physician’s first completed case assignment, and excluding completed replacement case assignments (Supplementary Table 5a). We also assessed how clustering by both physician and clinical case affected confidence intervals for the estimated differences between AI modes. A descriptive comparison included only points where AI advocated its instructed position. Across all entered case assignments, we examined whether physicians abandoned them more often with adversarial than aligned AI. Finally, we assessed how including final decisions recorded in incomplete or abandoned case assignments, or in completed case assignments by physicians who did not complete all four, could change the unadjusted harmful- and correct-decision differences between AI modes, under assumptions most favourable to adversarial AI and then to aligned AI (Supplementary Methods 11.8 and Supplementary Note 5).

In a sensitivity analysis, we calibrated the estimated effects on harmful and correct decisions and the proportions of harmful and correct-only AI advocacy using differences between independent human and LLM classifications. Approximate 95% uncertainty intervals accounted for sampling variability in both the full cohort and the human-reviewed samples, with standard errors clustered by physician (Supplementary Methods 10 and Supplementary Table 2).

All tests were two-sided, with Holm correction applied within defined secondary families; the primary harmful-decision test was unadjusted. Adjusted mode differences are reported with 95% confidence intervals without multiplicity adjustment. Analyses were conducted in Python (Supplementary Methods 11).

## Data availability

Full anonymized clinical cases will remain sequestered for future research. Requests for research access may be directed to the corresponding author. Adversarial instructions and operational details enabling their reuse will not be released publicly for safety reasons. Raw physician–AI conversations will remain non-public to protect participant privacy. De-identified decision-level classifications and analysis variables are not currently publicly available.

## Code availability

Analysis and figure code are not currently publicly available.

## Acknowledgements

We thank the Neurocore Research Group, the International Symposium on Intensive Care and Emergency Medicine (ISICEM), the Neurocritical Care Society (NCS), the Neurocritical Care and Neuro Anesthesiology French Speaking Society (ANARLF), and the Neurocritical Care Alliance for Research and Trials–Korea (NCART-KR) for their support with participant recruitment.

## Author contributions

**Sami Barrit**: Conceptualization; data curation; formal analysis; funding acquisition; investigation; methodology; project administration; resources; software; supervision; validation; visualization; writing – original draft; writing – review & editing. **Michele Salvagno**: Data curation; formal analysis; investigation; methodology; project administration; resources; writing – original draft; writing – review & editing. **Alberto Corriero** and **Trinity Pate**: Data curation; formal analysis; investigation; validation; writing – review & editing. **Madhumita Sushil**: Methodology; writing – review & editing. **Co-investigators** (JK, RS, MA, BB, SB, NE, NBH, RC, MB, GC, FH, CR, DV, MM, JCH, BF, RH, CL, DP, MK, SP): Data curation;\ investigation; writing – review & editing. **Neurocore research group**: Data curation; writing – review & editing. **Fabio Silvio Taccone** and **Edward F. Chang**: Funding acquisition; methodology; project administration; resources; supervision (lead); validation; writing – review & editing.

## Full authorship and affiliations

### Lead investigators

Sami Barrit, MD^1,2^; Edward F. Chang, MD^1^

*Department of Neurological Surgery*^1^ *and Computational Precision Health*^2^*, University of California, San Francisco and Berkeley, United States*

Michele Salvagno, MD; Fabio Silvio Taccone, MD, PhD

*Department of Intensive Care, Hôpital Universitaire de Bruxelles, Université Libre de Bruxelles, Belgium*

### Co-investigators

Marcel Aries, MD, PhD

*Department of Intensive Care Medicine, Universiteit Maastricht, Netherlands*

Marta Baggiani, MD

*Department of Neurocritical Care, Fondazione Istituto di Ricovero e Cura a Carattere Scientifico San Gerardo dei Tintori, Monza, Italy*

Baptiste Balança, MD, PhD

*Department of Anesthesiology and Intensive Care, Hospices civils de Lyon, France; Université de Lyon, France*

Nawfel Ben-Hamouda, MD, MSc

*Department of Intensive Care Medicine, Centre hospitalier universitaire vaudois and Université de Lausanne, Switzerland*

Sarah Benghanem, MD

*Department of Intensive Care Medicine, Assistance publique – Hôpitaux de Paris, France*

Grégoire Cane, MD

*Department of Anesthesiology and Intensive Care, Centre hospitalier universitaire de Bordeaux, France*

Russell Chabanne, MD, MSc

*Department of Anesthesiology and Intensive Care, Université Clermont Auvergne, Clermont-Ferrand, France*

Alberto Corriero, MD, PhD

*Department of Interdisciplinary Medicine, Università degli Studi di Bari Aldo Moro, Italy*

Nicolas Engrand, MD, MSc

*Department of Neurocritical Care, Hôpital Fondation Adolphe de Rothschild, Paris, France*

Brandon Foreman, MD

*Department of Neurology and Rehabilitation Medicine, University of Cincinnati, United States*

Raimund Helbok, MD

*Department of Neurology, Johannes Kepler Universität Linz, Austria*

J. Claude Hemphill, MD

*Department of Neurology, University of California, San Francisco, United States*

Floriane Hilaire, MD

*Department of Anesthesiology and Intensive Care, Université de Bordeaux, France*

Matthew Kirschen, MD, PhD

*Department of Anesthesiology and Critical Care Medicine, University of Pennsylvania, Philadelphia, United States*

Julian Klug, MD

*Department of Intensive Care Medicine, Centre hospitalier universitaire vaudois, Lausanne, Switzerland*

Christos Lazaridis, MD

*Department of Neurology, The University of Chicago, United States*

Marina Munari, MD

*Department of Anesthesiology and Intensive Care, Azienda Ospedale-Università Padova and Università degli Studi di Padova, Italy*

Soojin Park, MD

*Department of Neurology, Columbia University, New York, United States*

Daniel Pinggera, MD, PhD

*Department of Neurosurgery, Medizinische Universität Innsbruck, Austria*

Chiara Robba, PhD

*Department of Anesthesiology and Intensive Care, Università degli Studi di Genova, Italy*

Rossana Soloperto, MD

*Department of Intensive Care, Hôpital Universitaire de Bruxelles, Université Libre de Bruxelles, Belgium*

Madhumita Sushil, PhD^1,2^

*Department of Neurological Surgery*^1^ *and Computational Precision Health*^2^*, University of California, San Francisco and Berkeley, United States*

Dmitar Vlahović, MD

*Department of Emergency Neurology, Klinika za neurologiju, Univerzitetski klinički centar Vojvodine, Novi Sad, Serbia*

### Contributors

Paer-Sélim Abback, MD

*Department of Anesthesiology and Critical Care Medicine, Centre hospitalier universitaire de Tours, France*

Leila Abbas, MD^1^; Clémence Marois, MD^2^; Nicolas Weiss, MD, PhD^2,3^

*Sorbonne Public Health Institute*^1^*, Sorbonne Université and Hôpital Pitié-Salpêtrière; Department of Neurocritical Care*^2^*, Hôpital Pitié-Salpêtrière; Sorbonne Université*^3^*, Paris, France*

Amit Agrawal, MCh

*Department of Neurosurgery, All India Institute of Medical Sciences Bhopal, India*

Cecilia Åkerlund, MD, PhD; Linn Hallqvist, MD, PhD

*Department of Physiology and Pharmacology, Karolinska Institutet, Stockholm, Sweden*

Mejdeddine Al Barajraji, MD^1^; Lorenzo Gavino, MD^2^

*Departments of Neurosurgery*^1^ *and Critical Care Medicine*^2^*, Centre hospitalier universitaire vaudois, Lausanne, Switzerland*

Daniela Alampi, MD^1^; Luigi Valentino Berra, MD^2^; Antonio Lauletta, MD, PhD^3^

*Departments of Medical-Surgical Sciences and Translational Medicine*^1^*, Neurosurgery*^2^ *and Neuroscience, Mental Health and Sensory Organs*^3^*, Azienda Ospedaliero-Universitaria Policlinico Umberto I and Sapienza Università di Roma, Italy*

Maxime Théo Aparicio, MD

*Department of Anesthesiology and Surgical Intensive Care, Hôpital européen Georges-Pompidou, Paris, France*

Raffaele Aspide, MD

*Department of Neurocritical Care, Istituto delle Scienze Neurologiche di Bologna, Italy*

Denise Battaglini, MD, PhD

*Department of Surgical Sciences and Integrated Diagnostics, Università degli Studi di Genova, Italy*

Hemant Bhagat, MD, DM^1^; Manjul Tripathi, MCh^2^

*Departments of Neuroanesthesiology and Neurocritical Care*^1^ *and Neurosurgery*^2^*, Postgraduate Institute of Medical Education and Research, Chandigarh, India*

Daniele Guerino Biasucci, MD, MSc^1^; Domenico Junior Brunetti, MD, PhD^2^; Francesco M.C. Lioi, MD^3^

*Departments of Clinical Science and Translational Medicine*^1^*, Intensive Care Medicine*^2^ *and Neurosurgery*^3^*, Fondazione Policlinico Tor Vergata and Università degli Studi di Roma “Tor Vergata”, Italy*

Summit Bloria, DM

*Department of Anesthesiology, Shri Mata Vaishno Devi Institute of Medical Excellence, Jammu, India*

Stefan Yu Bogli, MD^1^; Victor E. Staartjes, MD, PhD^2^

*Departments of Neurology*^1^ *and Neurosurgery*^2^*, Universitätsspital Zürich, Switzerland*

Marco Bonato, MD^1^; Tommaso Pettenuzzo, MD^2^; Silvia Piovesan, MD^2^

*Departments of Anesthesiology, Intensive Care and Pain Medicine*^1^ *and Medicine*^2^*, Azienda Ospedale-Università Padova and Università degli Studi di Padova, Italy*

Robert Hitchens Bonow, MD^1^; Abhijit Vijay Lele, MD^2^

*Departments of Neurosurgery*^1^ *and Anesthesiology, Neurology and Neurosurgery*^2^*, University of Washington, Seattle, United States*

Frederico Bruzzi De Carvalho, MD, MSc

*Department of Intensive Care and Burns, Hospital de Pronto-Socorro João XXIII, Belo Horizonte, Brazil*

Barbara Buccilli, MD

*Department of Neurosurgery and Engineering, The University of Oklahoma, Oklahoma City, United States*

Joseph D Burns, MD

*Department of Neurology, Lahey Hospital & Medical Center and University of Massachusetts Chan Medical School, Burlington, United States*

Katharina Maria Busl, MD, MS; Ivan Da Silva, MD, PhD

*Department of Neurology, University of Florida, Gainesville, United States*

Anna Cervantes-Arslanian, MD

*Department of Neurology, Boston Medical Center and Boston University Chobanian & Avedisian School of Medicine, United States*

Viviana Chediack, MD; Eleonora Roxana Cunto, MD

*Department of Intensive Care Medicine, Hospital de Infecciosas Francisco Javier Muñiz, Buenos Aires, Argentina*

Sorayouth Chumnanvej, MD, PhD

*Department of Surgery, Faculty of Medicine Ramathibodi Hospital, Mahidol University, Bangkok, Thailand*

Elia Citton, MD

*Department of Critical Care Medicine, Imperial College Healthcare National Health Service Trust, London, United Kingdom*

Serena Crisci, MD

*Department of Anesthesiology, Intensive Care and Pain Medicine, Fondazione Poliambulanza Istituto Ospedaliero, Brescia, Italy*

Patrick Czorlich, MD^1^; Jamil Dibu, MD^2^; Massimo Lamperti, MD^3^

*Departments of Critical Care Medicine*^1^*, Neurocritical Care*^2^ *and Anesthesiology*^3^*, Cleveland Clinic Abu Dhabi, United Arab Emirates*

Airton Leonardo De Oliveira Manoel, MD, PhD^1^; Ali Msheik, MD^2^

*Departments of Trauma Surgery*^1^ *and Neurosurgery*^2^*, Hamad Medical Corporation, Doha, Qatar*

Pia De Stefano, MD

*Department of Neurology, Hôpitaux universitaires de Genève, Switzerland*

Greet De Vlieger, MD, PhD; Jan Gunst, MD, PhD

*Department of Intensive Care Medicine, Universitair Ziekenhuis Leuven and Katholieke Universiteit Leuven, Belgium*

Charles Dehout, MD

*Department of Intensive Care Medicine, Hôpital Erasme, Brussels, Belgium*

Masoom Desai, MD, MSDS

*Department of Neurology, University of New Mexico, Albuquerque, United States*

Celeste Dias, MD, PhD; João Ferreira-Coimbra, MD, MSc

*Department of Intensive Care Medicine, Unidade Local de Saúde de São João, Porto, Portugal*

Konstantinos Dimitriadis, MD, MSc, MA, MBA; Moritz Luigi Schmidbauer, MD; Julia Zibold, MD

*Department of Neurology, Klinikum der Ludwig-Maximilians-Universität München, Germany*

Chiara Faso, MD^1^; Alberto Marabotti, MD^2^

*Departments of Anesthesiology and Intensive Care*^1^ *and Intensive Care Medicine*^2^*, Azienda Ospedaliero-Universitaria Careggi, Florence, Italy*

Katharina Feil, MD

*Department of Neurology, Universitätsklinikum Ulm, Germany*

Eberval Gadelha Figueiredo, MD, PhD

*Department of Neurosurgery, Universidade de São Paulo, Brazil*

Matteo Filippini, MD^1^; Carola Matellon, MD^2^

*Departments of Anesthesiology and Critical Care Medicine*^1^ *and Cardiothoracic Intensive Care*^2^*, Azienda Socio Sanitaria Territoriale degli Spedali Civili di Brescia, Italy*

Senta Frol, PhD

*Department of Vascular Neurology, Univerzitetni klinični center Ljubljana, Slovenia*

Peter Galos, MD, MSc

*Department of Medical Sciences, Uppsala universitet, Sweden*

Alberto Nicolò Galvano, MD

*Department of Anesthesiology and Intensive Care, Università degli Studi di Palermo, Italy*

Diederik Geboers, MD

*Department of Intensive Care Medicine, Amsterdam Universitair Medische Centra, Netherlands*

Fabrizio Giammello, MD, PhD

*Department of Neurology and Stroke, Réseau hospitalier neuchâtelois, Neuchâtel, Switzerland*

Andres Giglio, MD, MSc, MHA, MPH

*Department of Medicine, Universidad Finis Terrae, Santiago, Chile*

Nicolas Higel, MD

*Department of Intensive Care Medicine, Centre hospitalier universitaire de Guyane, Cayenne, French Guiana*

Christopher Hohmann, MD

*Department of Internal Medicine, Medizinische Fakultät und Uniklinik Köln, Universität zu Köln, Germany*

Patrick M Honore, MD, PhD, FCCM

*Department of Intensive Care Medicine, Centre hospitalier universitaire Université catholique de Louvain Namur, Yvoir, Belgium*

Jaeho Hwang, MD, MPH

*Department of Neurology, Northwestern University, Chicago, United States*

Neel D. Jani, MD

*Department of Neurology, University of California, San Francisco, United States*

Rafael Batista João, MSc

*Department of Neurology, Universidade Federal de Goiás, Goiânia, Brazil*

John Kanter, MD^1^; Alexis Steinberg, MS^2^

*Departments of Neurosurgery and Critical Care Medicine*^1^ *and Neurology*^2^*, University of Pittsburgh Medical Center and University of Pittsburgh, United States*

Indu Kapoor, MD, PhD; Charu Mahajan, MD, DM; Hemanshu Prabhakar, MD, PhD

*Department of Neuroanesthesiology and Neurocritical Care, All India Institute of Medical Sciences, New Delhi, India*

Thomas Kerz, MD

*Department of Anesthesiology, Universitätsmedizin der Johannes Gutenberg-Universität Mainz, Germany*

Daniel Kheradmand, MD

*Department of Neurosurgery, Mashhad University of Medical Sciences, Iran*

Ching Soong Khoo, MD, FRCP

*Department of Medicine, Universiti Kebangsaan Malaysia, Kuala Lumpur, Malaysia*

Ozlem Korkmaz Dilmen, MD

*Department of Anesthesiology and Intensive Care, İstanbul Üniversitesi-Cerrahpaşa Cerrahpaşa Tıp Fakültesi, Turkey*

Uttara Koul, MD

*Department of Neurocritical Care, St. Vincent’s Medical Center, Bridgeport, United States*

Karim Lakhal, MD

*Department of Anesthesiology and Critical Care Medicine, Centre hospitalier universitaire de Nantes, France*

Andrea Lavinio, MD, MBA, FCICM, FRCA, RCPathME

*Department of Perioperative, Acute, Critical Care and Emergency Medicine, University of Cambridge, United Kingdom*

Aaron Lawson Mclean, MD

*Department of Neurosurgery, Universitätsklinikum Jena, Germany*

Alexandre Ledos, MD, MSc

*Department of Surgical Intensive Care, Centre hospitalier universitaire de Rennes, France*

Stephane Legriel, MD, PhD

*Department of Intensive Care Medicine, Centre hospitalier de Versailles, France*

Andrea Loggini, MD, MPH, MBA

*Department of Neurology, Southern Illinois Healthcare, Carbondale, United States*

Rossana Geraldine López, MD, MSc

*Department of Intensive Care Medicine, Clínica Privada Pueyrredón, Buenos Aires, Argentina*

Brandon Lucke-Wold, MD, PhD

*Department of Neurosurgery, University of Arizona, Tucson, United States*

Takuma Maeda, MD, PhD

*Department of Neurosurgery, Saitama Medical University International Medical Center, Hidaka, Japan*

Ali Mahta, MD, MA, FAHA, FAAN, FNCS

*Department of Neurosciences, University of California, San Diego Medical Center, United States*

Nicoló Marchesini, MD

*Department of Neurosurgery, Azienda Ospedaliera Universitaria Integrata di Verona, Italy*

Alexis Marouk, MD, MSc^1^; Roman Mounier, MD, PhD^2^

*Departments of Emergency Medicine*^1^ *and Anesthesiology and Critical Care Medicine*^2^*, Hôpital Avicenne, Assistance publique – Hôpitaux de Paris, Bobigny, France*

Nina Massad, MD^1^; Wander Leao Valentim, MD^2^

*Departments of Neurology and Neurocritical Care*^1^ *and Neurology*^2^*, University of Miami, United States*

Victoria McCredie, MBChB, PhD

*Department of Critical Care Medicine, University Health Network, Toronto, Canada*

Hamida Memon, MBBS

*Department of Medicine, Dow University of Health Sciences, Karachi, Pakistan*

Mostafa Meshref, MD, MSc, PhD

*Department of Neurology, Faculty of Medicine, Al-Azhar University, Cairo, Egypt*

Emanuel Moisa, MD, PhD, DESAIC

*Department of Anesthesiology and Critical Care Medicine, Universitatea de Medicină și Farmacie „Carol Davila” din București, Romania*

Luis Rafael Moscote-Salazar, MD

*Department of Research, AV Healthcare Innovators, LLC, Madison, United States*

Dilmareth E. Natera, MD

*Department of Neurosurgery, University of Minnesota, Minneapolis, United States*

Anthony Nguyen, MD

*Department of Neurosurgery, Baylor Scott & White Health, Temple, United States*

Nhan Nguyen, MD

*Department of Pediatrics, State University of New York Upstate Medical University, Syracuse, United States*

Alexandre Niset, MD

*Department of Emergency Medicine, Cliniques universitaires Saint-Luc, Brussels, Belgium*

Elhadjene Nory, MD, MSc

*Department of Anesthesiology and Intensive Care, Assistance publique – Hôpitaux de Marseille, France*

Yusuke Otsu, MD

*Department of Neurosurgery, Kurume University, Japan*

Manoj Padmanabhan, DO

*Department of Neurosurgery, University of Michigan, Ann Arbor, United States*

Trinity Pate, MSc

*Neurocore, Consciense Foundation, London, United Kingdom*

Nathalia Elena Penagos Vargas, MD, MSc

*Department of Pediatric Neurology, Fundación Hospital Pediátrico la Misericordia, Bogotá, Colombia*

David Pérez-Torres, MD

*Department of Intensive Care Medicine, Hospital Universitario Río Hortega, Valladolid, Spain*

Edoardo Picetti, MD

*Department of Anesthesiology and Intensive Care, Azienda Ospedaliero-Universitaria di Parma, Italy*

Jamir Pitton Rissardo, MD

*Department of Neurology, Cooper University Hospital, Camden, United States*

Manoj Kumar R, MD; Vijay Sundarsingh, MD

*Department of Critical Care Medicine, Father Muller Medical College, Mangaluru, India*

Michele Romoli, MD, PhD

*Department of Neurology and Stroke, Ospedale Maurizio Bufalini, Cesena, Italy*

Clemens M Schirmer, MD, PhD, MBA

*Department of Neurosurgery, Geisinger, Wilkes-Barre, United States*

Marcus Josephus Schultz, MD, PhD

*Department of Anesthesiology, Kantonsspital St. Gallen, Switzerland*

Nathan A. Shlobin, MD, MBA

*Department of Neurology, Westchester Medical Center, Valhalla, United States*

Andre Siahaan, MD, PhD

*Department of Neurosurgery, Universitas Sumatera Utara, Medan, Indonesia*

Stein Silva, MD, PhD

*Department of Intensive Care Medicine, Centre hospitalier universitaire de Toulouse, France*

Ashwin Subramaniam, MBBS, GChPOM, MMed, FRACP, FCICM, PhD

*Department of Intensive Care Medicine, Dandenong Hospital, Monash Health, Melbourne, Australia*

Jotaro Tachino, MD, PhD

*Department of Traumatology and Acute Critical Care Medicine, The University of Osaka, Japan*

Shahed Toossi, MD

*Department of Neurology and Neurosurgery, Dell Medical School, The University of Texas at Austin, United States*

Nathan Torcida, MD

*Department of Neurology, Hôpital Universitaire de Bruxelles, Belgium*

Panayiotis N Varelas, MD, PhD

*Department of Neurology, Albany Medical College, United States*

Daniel Vázquez Justes, PhD

*Department of Neurology, Hospital Universitari Arnau de Vilanova, Lleida, Spain*

Eva Vitali, MD

*Department of Anesthesiology and Intensive Care, Università Politecnica delle Marche, Ancona, Italy*

Charikleia S. Vrettou, MD, PhD

*Department of Intensive Care Medicine, School of Medicine, National and Kapodistrian University of Athens, Greece*

Johannes Weller, MD

*Department of Neuroimmunology, Universitätsklinikum Bonn, Germany*

Lucas O Woehl, MSc

*Department of Medicine, Universidade do Planalto Catarinense, Lages, Brazil*

Luigi Zattera, MD

*Department of Anesthesiology and Intensive Care, Hospital Clínic de Barcelona, Spain*

Frederick A. Zeiler, MD, PhD, FRCSC

*Department of Surgery, University of Manitoba, Winnipeg, Canada*

